# Association between surgical aortic valve replacement and long-term outcomes in 50 to 65-year-olds: Results of the AUTHEARTVISIT study

**DOI:** 10.1101/2024.12.19.24319314

**Authors:** A. Florian, J. Auer, B. Reichardt, P. Krotka, C. Wagenlechner, R. Wendt, M. Mildner, J. Mascherbauer, HJ Ankersmit, D. Zimpfer, A. Graf

**Affiliations:** Department of Cardiac Surgery, Medical University of Vienna, Austria; Department of Internal Medicine I with Cardiology and Intensive Care, St. Josef Hospital Braunau, Braunau am Inn, Austria; Austrian Social Health Insurance Fund, Eisenstadt, Austria; Center for Medical Data Science, Medical University of Vienna, Austria; Department of Nephrology, St. Georg Hospital, Leipzig, Germany; Department of Dermatology, Medical University of Vienna, Austria; Department of Internal Medicine 3, University Hospital St. Poelten, St. Poelten, Austria; Karl Landsteiner University of Health Sciences, Krems an der Donau, Austria; Department of Thoracic Surgery, Medical University of Vienna, Austria; Laboratory for Cardiac and Thoracic Diagnosis, Regeneration and Applied Immunology, Austria

## Abstract

**Objectives:** During the last years, age recommendations for the use of biological prostheses rather than mechanical prostheses for surgical aortic valve replacement (sAVR) have been lowered considerably. We evaluated survival rates, major adverse cardiac events (MACEs), and reoperation risks after surgical (sM-AVR) and biological (sB-AVR) AVR, to provide data for the optimal prosthesis choice for middle aged patients between 50 and 65 years.

**Methods:** We performed a population-based cohort study using Austrian Health System data from 2010–2020. Patients undergoing isolated sAVR (n=3761) were categorized into sM-AVR (n=1018) and sB-AVR (n=2743) groups. Propensity score matching (PSM) was applied to balance covariates. The primary endpoint was all-cause mortality. Secondary endpoints included MACEs, reoperation, stroke, bleeding, and survival post-reoperation. Outcomes were assessed using Cox regression and Kaplan-Meier analyses.

**Results:** Patients undergoing sM-AVR had significantly lower all-cause mortality compared to sB-AVR (HR=1.352, p=0.003). sB-AVR was associated with higher risks of MACEs (HR=1.182, p=0.03) and reoperation (HR=2.338, p=0.002). Stroke and bleeding rates were comparable. All results were sustained after PSM.

**Conclusion:** The findings highlight increased mortality, MACEs and reoperation risks associated with sB-AVR compared to sM-AVR. We observed superior long-term outcomes after sM-AVR, suggesting the need to reevaluate the expanding use of sB-AVR in younger patients.

**Graphical Abstract:** 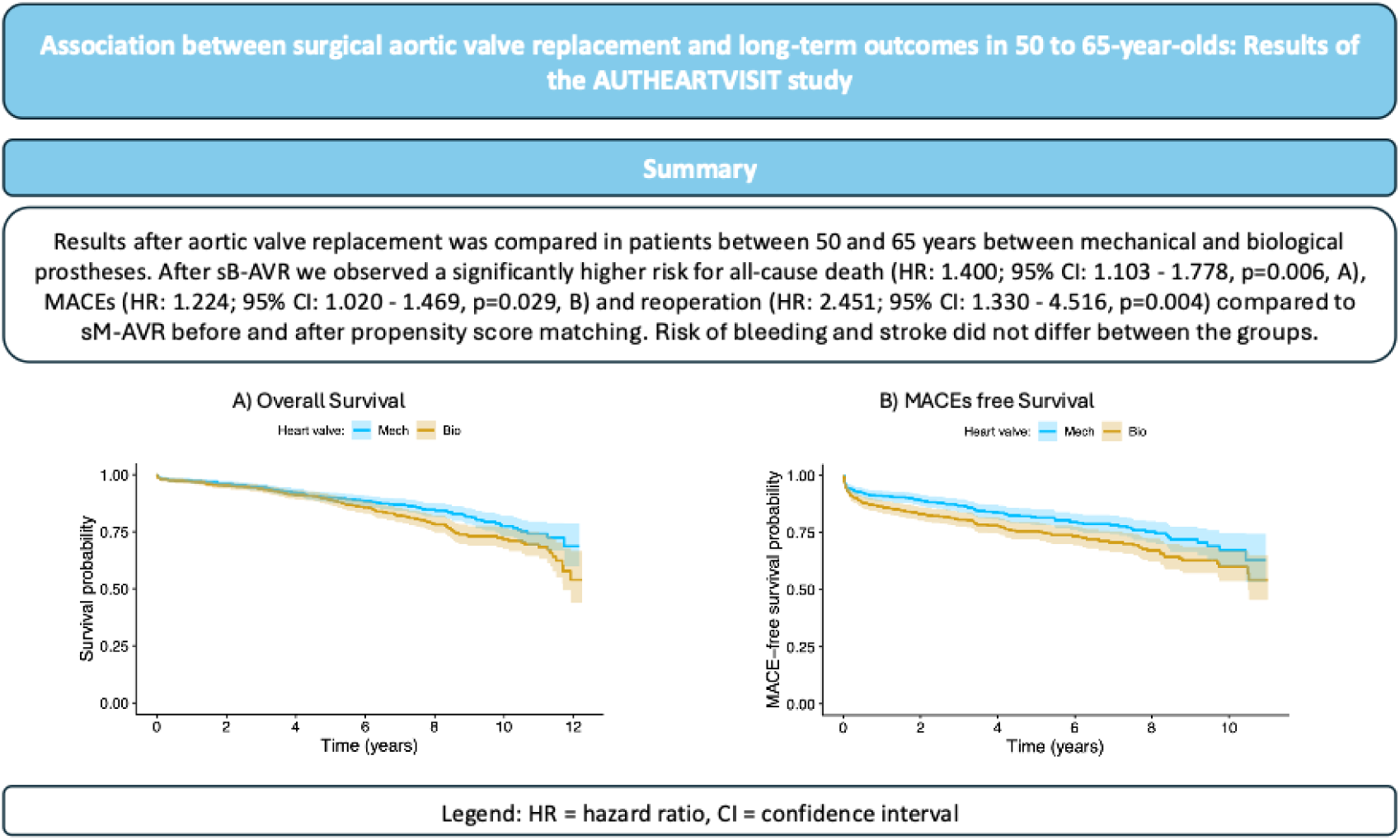

## Introduction

Valvular heart disease is a leading global cause of cardiovascular morbidity and mortality with aortic valve disease accounting for approximately 61% of deaths from valvular heart disease. The prevalence of aortic valve stenosis varies by age, starting from 0.2% in 50 to 59-year-olds and reaching 9.8% in 80 to 89-year-olds (1,2). The standard treatment for severe aortic valve disease is aortic valve replacement (AVR) using biological and mechanical prostheses, homografts or autografts (3).

European Society of Cardiology (ESC) guidelines recommend mechanical prostheses for patients aged ≤60 years and biological prostheses for those ≥65 years. For patients aged 60-65, decisions depend on patient preferences and clinical factors (4). American Heart Association (AHA)/American College of Cardiology (ACC) guidelines suggest mechanical prostheses for patients ≤50 years, biological prostheses for those ≥65 years, and preference-based decisions for those aged 50-65 (5).

The lower age limits seem to be driven by some registry studies showing similar outcomes after both types of valve replacement. Similar long-term survival rates have been reported in middle aged patients (6,7) as well as in young patients under 50 years (8). Also a higher incidence of stroke was reported after sM-AVR (9). These results lead to more frequent use of bioprostheses in younger patients.

However, several studies have shown an increase in mortality, major adverse cardiac events (MACEs), and reoperations in association with sB-AVR (9–14). Our group published two papers regarding the choice of prosthesis. Besides finding that implantation of transcatheter aortic valves compared to surgical AVR in patients younger than 65 years of age is associated with higher all-cause mortality (15), Traxler et al. recently observed better long-term survival and a lower risk of reoperation after sM-AVR in patients aged 50-65 years (14) as well as in patients aged ≤50 years (13).

The described discrepancies show that the decision between sB-AVR and sM-AVR is not fully standardized and the optimal type of prosthesis for middle-aged patients (50-65 years of age) needs to be evaluated in more detail. Therefore, we conducted a population-based cohort study based on all patients aged between 50 and 65 years who underwent isolated surgical AVR in Austria between 2010 and 2020 and were covered by the main Austrian insurance carriers, which ensured their data were gathered. We extended our previously published study based on the AUTHEARTVISIT registry to gain stronger evidence by including additional patients who underwent surgery after 2018 and up to 2020 and performing propensity score matching (PSM) to create two comparable patient groups. Real-life data based on ICD-10, ATC, and MEL codes (see Supplementary Tables 1-5) were collected from the Austrian insurance funds. Our primary endpoint was the comparison of survival after sB-AVR or sM-AVR. Secondary endpoints were MACEs, reoperations, survival after reoperation, and stroke or bleeding after each procedure.

## Methods

### Study design

In compliance with the Declaration of Helsinki, we conducted a nationwide, population-based cohort study. The AUTHARTVISIT Study was approved by the ethics committee of lower Austria (GS1-EK-4/722-2021). The trial was registered at clinicaltrials.gov (NCT05912660). We obtained the clinical and operative data for all patients registered in the Austrian Health Care System who underwent surgical AVR (sM-AVR [MEL code DB082] or sB-AVR [MEL codes DB060, DB070, and DB080]) in Austria between the years 2010 and 2020 who were between 50 and 65 years of age (see Supplementary Tables 1 and 4).

Patients undergoing transcatheter aortic valve implantation (TAVI; MEL codes DB025, DB026, DB021, or XN010) as the index procedure were not included in the analyzed sample. Patients <50 years of age or >65 years of age were excluded from the study. Patients with concomitant heart surgery, including multivalvular surgery or additional procedures performed during the index operation, were also excluded.

Furthermore, patients receiving a coronary artery stent (MEL code DD050 “implantation of a stent in the coronary artery” or DD060 “implantation of a drug eluting stent in a coronary artery”) within 4 months prior to the AVR were excluded from this analysis to exclude patients with percutaneous coronary intervention and guarantee the selection of patients with pure AVR procedures (see Supplement Section 2). The data set included 3761 patients: 1018 who underwent sM-AVR and 2743 who underwent sB-AVR.

## Statistical analysis

Categorical variables are presented as counts and percentages. Continuous variables are summarized as the median and the 1st and 3rd quartile (interquartile range [IQR]). We primarily evaluated the association between prosthesis type (sB-AVR or sM-AVR) and all-cause death. As secondary outcomes, the time until MACEs, death or reoperation (reoperation-free survival), reoperation, heart failure, myocardial infarction, embolic stroke or ICH, and bleeding other than embolic stroke or ICH was investigated (see Supplementary Table 2). MACE is a combined endpoint consisting of death, reoperation, heart failure, heart attack, and embolic stroke or intracerebral hemorrhage (ICH). The exploratory outcome was time to all-cause death after reoperation.

The association between prosthesis type and the primary outcome, all-cause death, was evaluated using a Cox regression model. This model was adjusted for age, sex, and other co-morbidities present within 1 year before the index surgery, including myocardial Infarction; embolic stroke or ICH; diabetes mellitus; obesity; hyperlipidemia; hyperuricemia/gout; valvular, rhythmological, or other cardiomyopathies (CMPs); ischemic CMP; arteriosclerosis; pulmonary diseases; kidney diseases; and malignant diseases (see Supplementary Table 5). Kaplan-Meier curves were created and the results of the Cox regression model presented as hazard ratios (HRs) and corresponding 95% confidence intervals (CIs), as well as p-values. This model was repeated with the interaction terms between sex and prosthesis type, as well as age and prosthesis type. The proportional hazards assumption was evaluated using Schönfeld residuals, and collinearity was evaluated using variance inflation factors.

The secondary outcomes MACEs and death or reoperation were analyzed similar to the primary endpoint. The time until the remaining events (heart failure, myocardial infarction, embolic stroke or ICH, reoperation, bleeding other than embolic stroke or ICH) was assessed using competing risk regression including death as a competing event using the Fine and Grey method. Cumulative incidence functions were plotted and results of competing risk regression models presented as HRs and corresponding 95% CIs with p-values. Due to the small number of events, time to all-cause death after reoperation was only evaluated in a descriptive manner using the Kaplan-Meier method. HRs >1 for the predictor “prosthesis type” are to be interpreted as patients who underwent sB-AVR being at higher risk of experiencing the particular event compared to patients who underwent sM-AVR.

In sensitivity analyses, all models were also calculated for the subgroup of patients aged 50 - 60 years. Models for heart failure and MACEs were calculated for patients without heart failure before the index surgery. In additional sensitivity analyses, all models were repeated with the propensity score-matched prosthesis groups. Propensity scores were estimated by logistic regression based on the following cofounders: age, sex, and co-morbidities present 1 year before the index surgery (myocardial infarction; embolic stroke or ICH; diabetes mellitus; obesity; hyperlipidemia; hyperuricemia/gout; valvular, rhythmological, and other CMPs; ischemic CMP; arteriosclerosis; pulmonary diseases; kidney diseases; malignant diseases).

The matching was performed using the nearest neighbor method, 1:1 ratio, and caliper width of 0.01. Notably, after PSM, the sample size and event rates were lower; the full model including all confounders could not be calculated for all outcomes. Co-morbidities not included in the models are indicated as not available (NA).

All analyses were performed using R, version 4.3.2. All p-values < 0.05 were considered significant. Due to the retrospective and exploratory character of the study, no correction for multiplicity was applied. Therefore, results on secondary and exploratory outcomes should be interpreted with caution.

## Results

### Study population and patient characteristics

From 2010 to 2020, 3761 patients underwent surgical AVR: 1018 sM-AVR and 2743 sB-AVR. Over all observed years, more patients aged 50-65 years underwent sB-AVR, as did the subgroup of patients aged 50 – 60 years (Figure 1). Patients undergoing sB-AVR were generally older and included larger proportions of patients with diabetes mellitus, ischemic CMP, and malignant diseases, whereas a significantly larger proportion of patients who underwent sM-AVR had hyperuricemia/gout (see Table 1). For detailed information on prescribed medicaments, see Supplementary Table 6.

**Figure 1:**
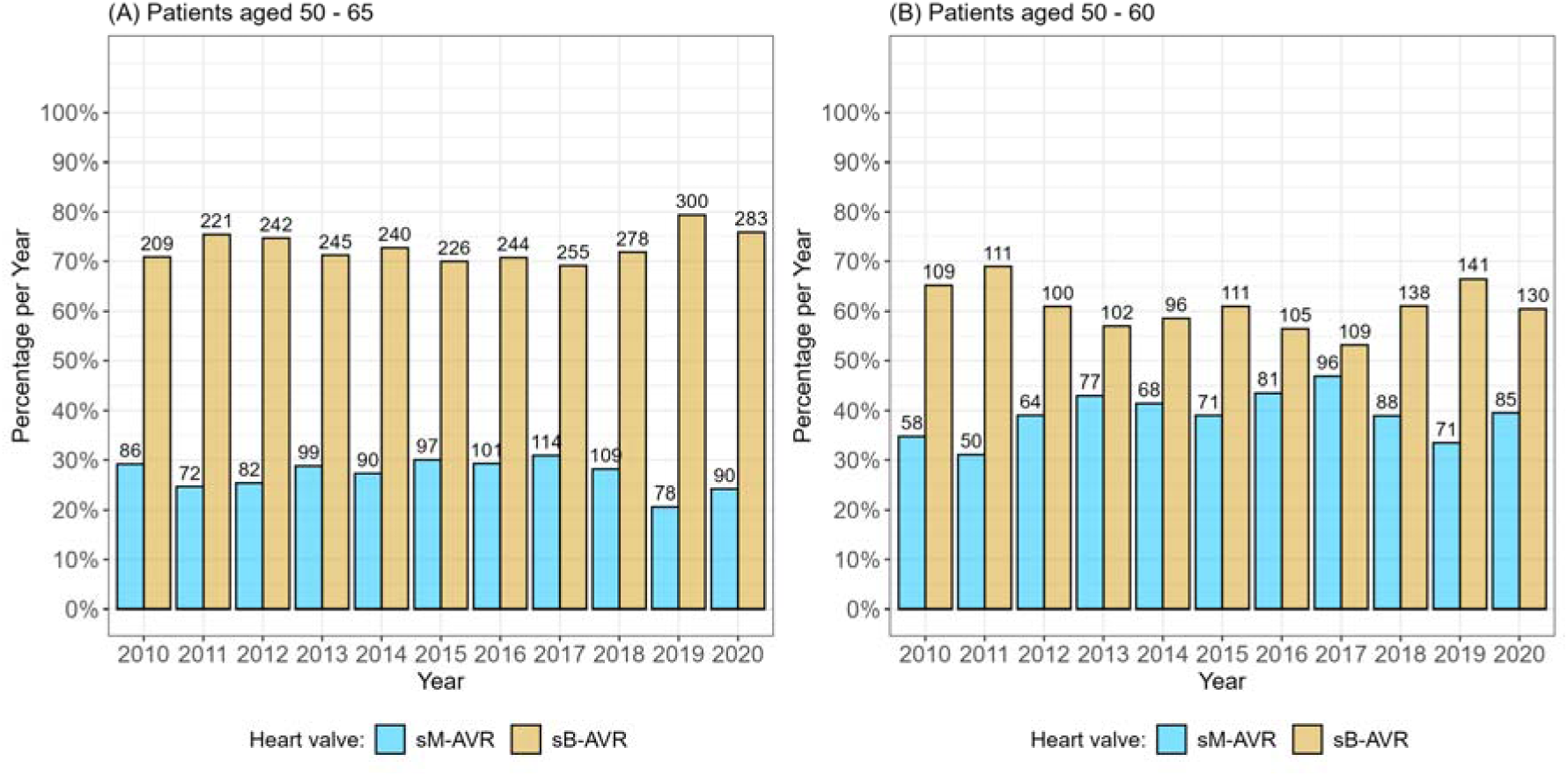
Percentages per year (y-axis) and absolute numbers (above the bars) of patients receiving biological or mechanical valves. (A) Patients aged 50 – 65 years. (B) The subgroup of patients aged 50 - 60 years.

**Table 1:** Characteristics and pre-existing medical diagnoses at the time of the index operation. Unless otherwise indicated, the values are given as the number of patients with a diagnosis and the corresponding percentage of total patients in the treatment group.

|  | All data |  |  | After PSM |  |  |
| --- | --- | --- | --- | --- | --- | --- |
| Variables | sM-AVR<br>(n=1018) | sB-AVR<br>(n= 2743) | P-value | sM-AVR<br>(n=902) | sB-AVR<br>(n= 902) | P-value |
| **Patients aged 50 – 65 years** |  |  |  |  |  |  |
| Median age, years (IQR) | 56 (53 - 60) | 61 (57 - 63) | <0.001 | 57 (54 - 60) | 57 (54 - 60) | 0.913 |
| Sex, female | 284 (27.9%) | 743 (27.09%) | 0.649 | 255 (28.27%) | 233 (25.83%) | 0.266 |
| Heart failure | 91 (8.94%) | 295 (10.75%) | 0.116 | 74 (8.2%) | 78 (8.65%) | 0.799 |
| Myocardial infarction | 24 (2.36%) | 93 (3.39%) | 0.130 | 23 (2.55%) | 18 (2%) | 0.527 |
| Embolic stroke or ICH | 9 (0.88%) | 48 (1.75%) | 0.075 | 9 (1%) | 8 (0.89%) | 1 |
| Diabetes mellitus | 118 (11.59%) | 441 (16.08%) | <0.001 | 111 (12.31%) | 105 (11.64%) | 0.717 |
| Obesity | 84 (8.25%) | 233 (8.49%) | 0.863 | 71 (7.87%) | 62 (6.87%) | 0.471 |
| Hyperlipidemia | 254 (24.95%) | 647 (23.59%) | 0.408 | 207 (22.95%) | 205 (22.73%) | 0.955 |
| Hyperuricemia/gout | 40 (3.93%) | 71 (2.59%) | 0.040 | 27 (2.99%) | 28 (3.1%) | 1 |
| Valvular, rhythmological, and other CMPs | 839 (82.42%) | 2307 (84.1%) | 0.232 | 739 (81.93%) | 735 (81.49%) | 0.855 |
| Ischemic CMP | 394 (38.7%) | 1252 (45.64%) | <0.001 | 340 (37.69%) | 357 (39.58%) | 0.439 |
| Arteriosclerosis | 33 (3.24%) | 119 (4.34%) | 0.154 | 31 (3.44%) | 20 (2.22%) | 0.155 |
| Pulmonary diseases | 31 (3.05%) | 102 (3.72%) | 0.371 | 27 (2.99%) | 25 (2.77%) | 0.888 |
| Kidney diseases | 65 (6.39%) | 194 (7.07%) | 0.505 | 57 (6.32%) | 51 (5.65%) | 0.620 |
| Malignant diseases | 16 (1.57%) | 85 (3.1%) | 0.014 | 13 (1.44%) | 15 (1.66%) | 0.849 |
| **Patients aged 50 – 60 years** |  |  |  |  |  |  |
| Median age, years (IQR) | 55 (53 - 57) | 57 (54 - 59) | <0.001 | 56 (53 - 58) | 55 (53 - 58) | 0.775 |
| Sex, female | 219 (27.07%) | 290 (23.16%) | 0.050 | 171 (25.37%) | 172 (25.52%) | 1 |
| Heart failure | 61 (7.54%) | 110 (8.79%) | 0.358 | 39 (5.79%) | 47 (6.97%) | 0.435 |
| Myocardial infarction | 16 (1.98%) | 36 (2.88%) | 0.261 | 12 (1.78%) | 9 (1.34%) | 0.660 |
| Embolic stroke or ICH | 6 (0.74%) | 26 (2.08%) | 0.027 | 5 (0.74%) | 4 (0.59%) | 1 |
| Diabetes mellitus | 76 (9.39%) | 161 (12.86%) | 0.019 | 58 (8.61%) | 62 (9.2%) | 0.774 |
| Obesity | 65 (8.03%) | 99 (7.91%) | 0.983 | 43 (6.38%) | 42 (6.23%) | 1 |
| Hyperlipidemia | 194 (23.98%) | 265 (21.17%) | 0.148 | 150 (22.26%) | 132 (19.58%) | 0.255 |
| Hyperuricemia/gout | 28 (3.46%) | 24 (1.92%) | 0.041 | 16 (2.37%) | 12 (1.78%) | 0.567 |
| Valvular, rhythmological, and other CMPs | 665 (82.2%) | 1042 (83.23%) | 0.587 | 536 (79.53%) | 546 (81.01%) | 0.538 |
| Ischemic CMP | 292 (36.09%) | 492 (39.3%) | 0.157 | 250 (37.09%) | 241 (35.76%) | 0.651 |
| Arteriosclerosis | 25 (3.09%) | 40 (3.19%) | 0.997 | 20 (2.97%) | 18 (2.67%) | 0.869 |
| Pulmonary diseases | 20 (2.47%) | 42 (3.35%) | 0.311 | 15 (2.23%) | 17 (2.52%) | 0.858 |
| Kidney diseases | 46 (5.69%) | 73 (5.83%) | 0.967 | 38 (5.64%) | 30 (4.45%) | 0.384 |
| Malignant diseases | 11 (1.36%) | 34 (2.72%) | 0.057 | 10 (1.48%) | 4 (0.59%) | 0.179 |

Due to this imbalance between groups, we performed PSM (see Table 1).

### Primary outcome

For sB-AVR, we observed a significantly higher risk of all-cause death compared to sM-AVR (HR: 1.352; 95% CI: 1.109 - 1.649, p=0.003, Figure 2 and 3A). This was also observed in the subgroup of patients aged 50 – 60 years (HR: 1.601; 95% CI: 1.235 - 2.074, p<0.001, Figure 2 and 3C). PSM did not change the result in the overall cohort (HR: 1.400; 95% CI: 1.103 - 1.778, p=0.006, Figure 2 and 3C) or the subgroup aged 50 – 60 years (HR: 1.645; 95% CI: 1.209 - 2.238, p=0.002, Figure 2 and 3D).

**Figure 2:**
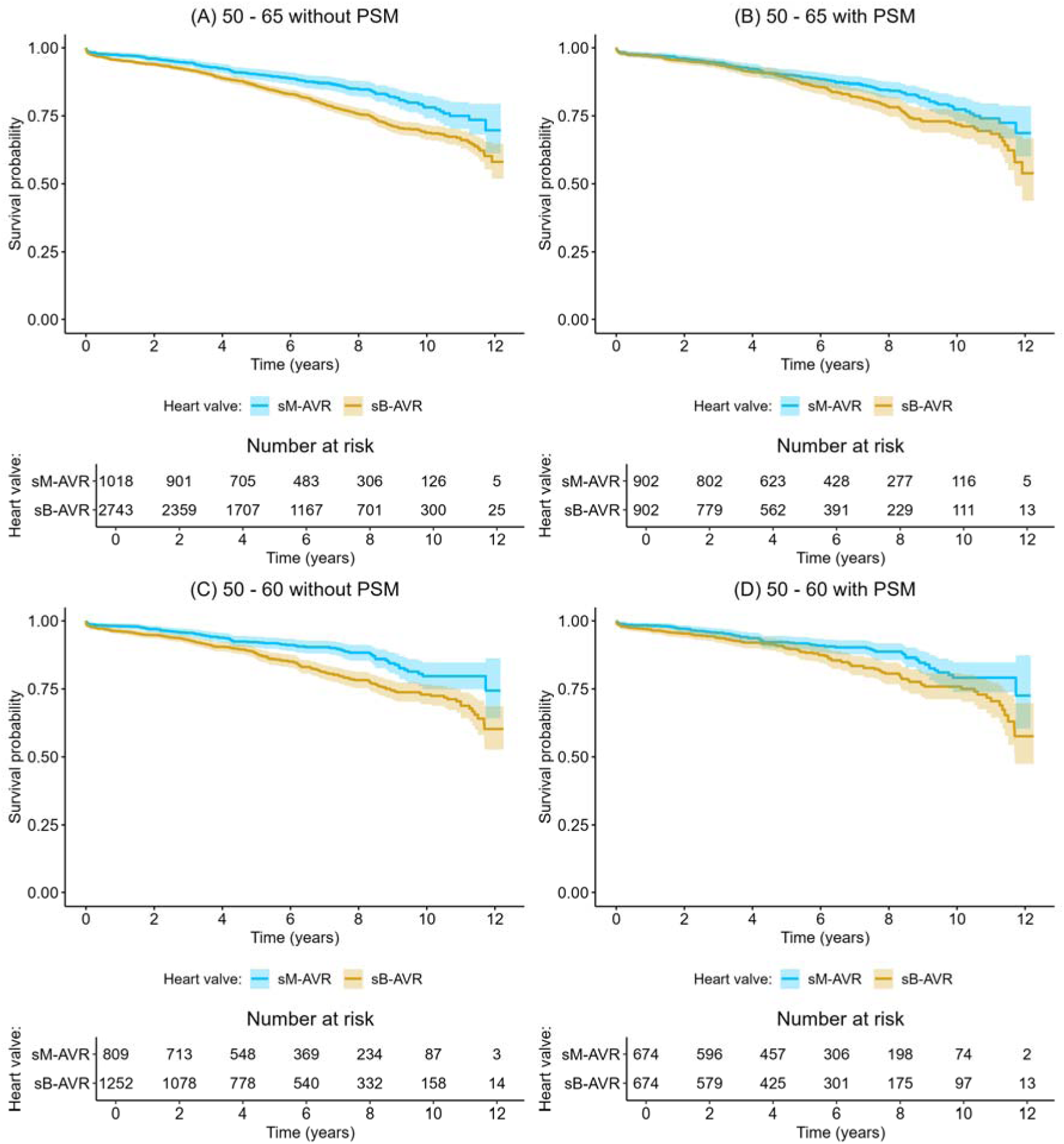
Kaplan-Meier curves and 95% confidence intervals for all-cause death before (A,C) and after (B,D) PSM for all patients aged 50 – 65 years (A,B) and the subgroup of patients aged 50 – 60 years (C,D)

**Figure 3:**
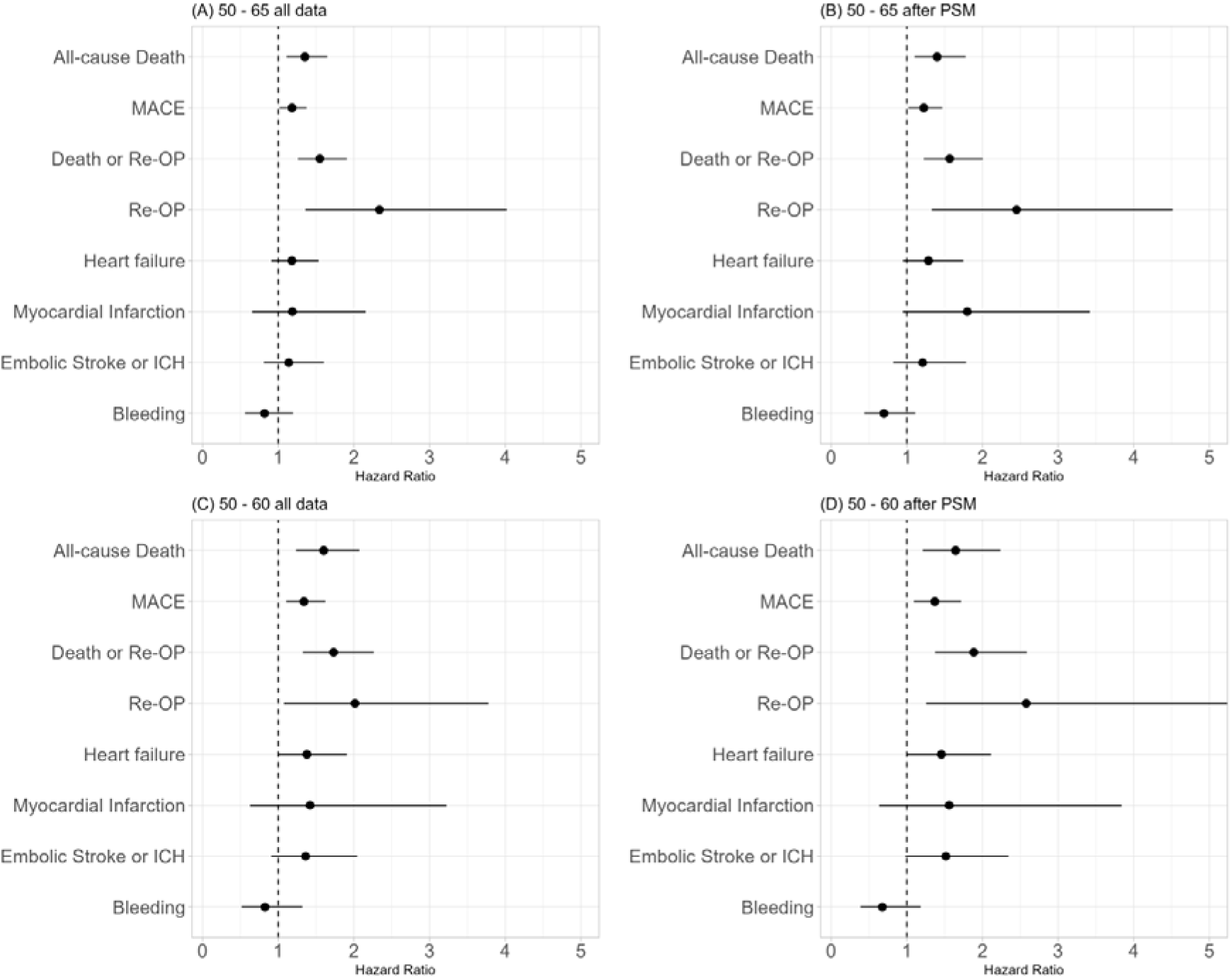
Hazard ratios (HRs) and 95% confidence intervals from multivariable regression models for all outcomes before (A, C) and after (B,D) PSM for all patients aged 50 – 65 years (A,B) and the subgroup of patients aged 50 – 60 years (C,D). HR>1 indicates a higher risk of the event occurring with the biological valve than with the mechanical valve.

For further details on the statistical models, see Supplementary Table 7.

### Secondary outcomes

#### MACEs

A significantly higher risk of MACEs was observed for sB-AVR compared to sM-AVR (HR: 1.182; 95% CI: 1.016 - 1.375, p=0.03, Figure 3A). This was also observed in the subgroup of patients aged 50 – 60 years (HR: 1.340; 95% CI: 1.105 - 1.624, p=0.003, Figure 3C). PSM did not change the result in the overall cohort (HR: 1.224; 95% CI: 1.020 - 1.469, p=0.029, Figure 3B) or the subgroup aged 50 – 60 years (HR: 1.370; 95% CI: 1.092 - 1.718, p=0.007, Figure 3D). In the cohort of patients aged 50 – 65 years, we observed a trend towards a decreasing advantage of sM-AVR compared to sB-AVR concerning MACEs with age (p=0.042); however, after PSM, the interaction terms between prosthesis type and age, as well as sex, did not show significant results (see Supplementary Table 8, Figure S1).

### Reoperation-free survival

Furthermore, a significantly lower probability of reoperation-free survival (see Supplementary Table 9, Figure S2) was observed for sB-AVR compared to sM-AVR (HR: 1.550; 95% CI: 1.260 - 1.908, p<0.001, Figure 3A). This was also observed in the subgroup of patients aged 50 – 60 years (HR: 1.732; 95% CI: 1.325 - 2.264, p<0.001, Figure 3C). PSM did not change the result in the overall cohort (HR: 1.566; 95% CI: 1.223 - 2.005, p<0.001, Figure 3B) or the subgroup aged 50 – 60 years (HR: 1.885; 95% CI: 1.373 - 2.587, p<0.001, Figure 3D).

### Reoperation

Concerning reoperation as outcome (see Supplementary Table 10, Figure S3), a significantly higher risk of reoperation was observed for sB-AVR compared to sM-AVR (HR: 2.338; 95% CI: 1.360 - 4.019, p=0.002, Figure 3A). This was also observed in the subgroup of patients aged 50 – 60 years (HR: 2.015; 95% CI: 1.075 - 3.778, p=0.029, Figure 3C). PSM did not change the result in the overall cohort (HR: 2.451; 95% CI: 1.330 - 4.516, p=0.004, Figure 3B) or the subgroup aged 50 – 60 years (HR: 2.579; 95% CI: 1.255 - 5.298, p=0.01, Figure 3C). The interaction terms between prosthesis type and age, as well as sex, did not show significant results.

### Heart failure

We did not find a significant difference between sB-AVR and sM-AVR for the risk of newly diagnosed heart failure (Figure 3, Supplementary Table 11, Figure S4) in the overall cohort (p=0.21, after PSM: p=0.11). Although not significant, a trend of a larger risk of newly diagnosed heart failure was observed for sB-AVR within the subgroup of patients aged 50 – 60 years (HR: 1.379; 95% CI: 0.997 - 1.908, p=0.052). This was also observed after PSM (HR: 1.457; 95% CI: 1.005 - 2.114, p=0.047). The interaction terms between prosthesis type and age, as well as sex, did not show significant results.

### Myocardial infarction

We did not find a significant difference between sB-AVR and sM-AVR for the risk of myocardial infarction (Figure 3, Supplementary Table 12, Figure S5) in the overall cohort (p=0.57, after PSM: p=0.073) or the subgroup aged 50 – 60 years (p=0.40, after PSM: p=0.33).

### Embolic stroke and ICH

We also did not find a significant difference between sB-AVR and sM-AVR for the risk of embolic stroke or ICH (Figure 3, Supplementary Table 13, Figure S6) in the overall cohort (p=0.46, after PSM: p=0.34) or the subgroup aged 50 – 60 years (p=0.14, after PSM: p=0.062). Although the main effects were not significant in the overall cohort of patients aged 50 – 65 years, we observed a trend towards a decreasing advantage of sM-AVR compared to sB-AVR concerning embolic stroke or ICH with age (p=0.004). The same trend was observed in the subgroup of patients aged 50 – 60 years (p=0.029). After PSM, the interaction term between prosthesis type and age remained significant for the overall cohort (p=0.021) but not for the subgroup (p=0.35).

### Bleeding

We did not find a significant difference between sB-AVR and sM-AVR for the risk of bleeding other than embolic stroke or ICH (Figure 3, Supplementary Table 14, Figure S7) in the overall cohort (p=0.30, after PSM: p=0.13) or the subgroup aged 50 – 60 years (p=0.42, after PSM: p=0.17). Although the main effects were not significant, in the cohort of patients aged 50 – 65 years, we observed a trend towards an advantage of sM-AVR compared to sB-AVR concerning bleeding other than embolic stroke or ICH in females, whereas a disadvantage of sM-AVR was found for males (p=0.037). The same trend was observed in the subgroup of patients aged 50 – 60 years (p=0.045). After PSM, the interaction term between prosthesis type and age remained significant for the overall cohort (p=0.046) but not for the subgroup aged 50 – 60 years (p=0.14).

### Outcome after reoperation

For survival after reoperation, only a small number of events were observed. The 5-year survival rates were estimated to be 85.2% (95% CI: 68% - 100%) in the sM-AVR group and 75.8% (95% CI: 66.6% - 86.2%) in the sB-AVR group. In the subgroup of patients aged 50 – 60 years, the 5-year survival rates were estimated to be 90.9% (95% CI: 75.4% - 100%) in the sM-AVR group and 79.0% (95% CI: 66.6% - 93.6%) in the sB-AVR group (see Supplementary Figure S8).

## Discussion

We found that middle aged patients undergoing AVR in Austria had a significantly longer survival after sM-AVR compared to sB-AVR. PSM did not change this result. These results confirm previous findings showing that patients who underwent sB-AVR had a significantly higher 15-year-mortality (16).

Additionally, the risk of MACE and reoperation was significantly higher after sB-AVR. Stassano and colleagues found similar results with significantly higher rates of valve failure and reoperation after sB-AVR compared to sM-AVR (17).

Notably, ESC guidelines recommend sM-AVR for patients <60 years (4), but sB-AVR outnumbered sM-AVR overall (72.93% vs. 27.07%) and in patients aged 50–60 years (60.75% vs. 39.25%). We also observed, despite a Class IIb recommendation to consider the use of mechanical prostheses in patients already on long-time anticoagulation, that more than 9% of patients undergoing sB-AVR were treated with oral anticoagulants in the year prior to surgery (4.41% of patients received vitamin K antagonists and 4.7% received direct oral anticoagulants). AHA/ACC guidelines suggest sM-AVR only for those <50 years and claim a better hemodynamic status and lower thromboembolic risk sB-AVR (5), though our data did not confirm these advantages. In contrast to previous studies, there was no statistically significant difference in the incidence of bleeding events after sM-AVR. More specifically, we did not observe a significant increase in general bleeding or intracerebral bleeding, which we think may be of particular importance. Hammermeister and colleagues found that patients who underwent sB-AVR had a reduced risk of bleeding. No differences were found in systemic embolism, infective endocarditis, and valve thrombosis (16).

However, similar to previous studies (16), we found no statistically significant difference in the incidence of stroke. Furthermore, in the propensity score-matched cohort, we observed a significantly greater risk of reoperation after sB-AVR compared to sM-AVR. Notably, the survival probability 10 years after reoperation was 71% after sM-AVR and 62.7% after sB-AVR. However, the results on survival after reoperation are based on a reduced number of events.

Most of the additional information that we have regarding long-term survival after sB-AVR or sM-AVR comes from registry studies, such as the abovementioned studies by Schnittman (8) and Goldstone (9). Alex and colleagues observed no differences in survival, stroke, bleeding, or endocarditis but a higher incidence of reoperation 15 years after sB-AVR (18) and Glaser et al. showed better long-term survival and a lower risk of reoperation after sM-AVR (11). However, in contrast to our findings, a higher risk of major bleeding events was reported for the patient group with sM-AVR. One possible explanation is reporting bias in registry studies with underestimation of the total bleeding event rate. Vogt et al. found no difference in 5-year mortality or reoperation but a higher stroke rate in sM-AVR patients (3.3% vs. 1.5%) (19).

In four meta-analyses published between 2019 and 2024 by Diaz (20), Jiang (21), Warraich (22) and Leviner (23), sB-AVR was associated with significantly worse survival rates and higher incidences of reoperation alongside with lower rates of bleeding compared to sM-AVR.

Despite robust evidence of a survival benefit after sM-AVR compared to sB-AVR, over the past few years, guideline recommendations and current practice went in the opposite direction. The age limits decreased for sB-AVR rather than sM-AVR. Although sM-AVR has the benefit of longevity of the prostheses, sB-AVR may be preferred because it does not require lifelong anticoagulation therapy due to the promising but unproven concept of TAVI “valve-in-valve”-strategy as a less invasive option for future interventions in the case of bioprosthetic valve failure. This ongoing discussion led to our decision to up-scale the patient cohort from our previous study regarding the choice of aortic valve prosthesis to obtain an even more robust dataset. The current analysis include 3824 patients with a follow-up of up to 12 years and thus upgrades our previous cohort that included 2612 patients from the same age group with a significantly shorter follow-up. In addition, to further overcome the shortages of a retrospective analysis, we aimed to create two comparable groups by performing PSM and including stroke, ICH, and general bleeding in our analysis, as those are strong arguments in every discussion regarding prosthesis choice (14).

One of the downsides of registry studies and meta-analyses is that only existing data can be used, which is why almost exclusively survival, rate of reoperation, and MACEs are compared. Yet, there are studies showing further downsides of sB-AVR. Percy and his colleagues observed that subclinical structural valve degeneration (SVD) occurs in more than 40% of patients <65 years of age, 21.5% of whom progressed to clinical SVD or repeat aortic valve procedures. First signs of SVD occur as early as 11 months after sB-AVR (24). These results support the findings of Salaun et al. who found, after a median follow-up of 6.7 months, that 25.6% of patients had leaflet calcifications on computed tomography and 38% developed hemodynamic valve deterioration (25).

As many studies have observed age-dependent accelerated SVD after sB-AVR, which may lead to a higher incidence of reoperation, especially in young patients (26–28), and a possibly increased risk of death for patients undergoing reoperation, the choice of aortic valve prosthesis should be made with care. Also the consequence that younger patients may reject their bioprosthesis due to a humoral and cellular immune response should be included in the decision for the heart valve type (29–31).

## Limitations

Our study aimed to show real-world evidence through the utilization of collected data from the Austrian insurance funds. Like many health service research studies, the observational nature of our research comes with certain limitations. We have the great advantage of being able to collect data on all patients registered in the Austrian Health System who underwent sB-AVR or sM-AVR in Austria between 2010 and 2020. As health care in Austria is a national system with good access to care, the majority (∼98%) of the Austrian population is registered.

Although our data were collected retrospectively, meaning they do not adhere to the standards of a prospective randomized trial and controlled allocation of treatments was not performed, we aimed to create two comparable patient groups and reduce the possibility that sM-AVR was more frequently performed in younger, healthier individuals than sB-AVR by performing Propensity Score Matching.

Due to our study utilizing administrative data, the information comes from billing records and discharge codes. This means we are reliant on the accurate coding of diseases and events across the country for a dependable analysis. Unfortunately, we cannot verify or correct this data retrospectively, which could introduce bias and cause inconsistencies when compared to prospective clinical databases. Moreover, we were unable to thoroughly investigate the medical reasons behind deaths or reoperations among the patients in the dataset. As a result, it was not possible to exclude patients whose death or reoperation was unrelated to the original surgery. Nonetheless, the main outcome measures used in our study involve events that require hospital care, and these are likely to be reported accurately to the Austrian health insurance carriers for billing purposes.

## Conclusions

Based on our data from patients who underwent isolated AVR in Austria between 2010 and 2020, we suggest that the current trend in lowering the age limits for sB-AVR has to be re-evaluated critically. In our population-based study, we observed a significantly increased mortality and reoperation risk after sB-AVR in younger patients. Therefore, the choice of aortic valve prosthesis in younger patients should be handled with care.

## Data Availability

All data produced in the present study are available upon reasonable request to the authors

## Abbreviations

AVR: aortic valve replacement
CMP: cardiomyopathy
ICH: intracerebral hemorrhage
MACE: major adverse cardiac event
PSM: propensity score matching
sAVR: surgical aortic valve replacement
sB-AVR: biological aortic valve replacement
sM-AVR: mechanical aortic valve replacement
SVD: structural valve degeneration
TAVI: transcatheter aortic valve implantation

## Acknowledgements

We thank the Pharmaco-economics Advisory Council of the Austrian Sickness Funds for providing the data, especially Ms. Karin Allmer for quality assurance of the database query and Mr. Ludwig Weissengruber for organizational support for data generation.

