## Supplement for "Association between surgical aortic valve replacement and long-term outcomes in 50 to 65-year-olds: Results of the AUTHEARTVISIT study"

A. Florian^1^, J. Auer^2^, B. Reichardt^3^, P. Krotka^4^, C. Wagenlechner^4^, R. Wendt^5^, M. Mildner^6^, J. Mascherbauer^7,8^, HJ Ankersmit^9,10*#^, D. Zimpfer^1*#^, A. Graf^4#^

1 Department of Cardiac Surgery, Medical University of Vienna, Austria

2 Department of Internal Medicine I with Cardiology and Intensive Care, St. Josef Hospital Braunau, Braunau am Inn, Austria

3 Austrian Social Health Insurance Fund, Eisenstadt, Austria

4 Center for Medical Data Science, Medical University of Vienna, Austria

5 Department of Nephrology, St. Georg Hospital, Leipzig, Germany

6 Department of Dermatology, Medical University of Vienna, Austria

7 Department	of Internal Medicine 3, University Hospital St. Poelten, St. Poelten, Austria

8 Karl Landsteiner University	of Health Sciences, Krems an	der Donau, Austria

9 Department of Thoracic Surgery, Medical University of Vienna, Austria

10 Laboratory for Cardiac and Thoracic Diagnosis, Regeneration and Applied Immunology, Austria

\# contributed equally

* corresponding authors

1.) **General Information on Coding**

For each patient, billing information (based on MEL codes from the Austrian insurance carriers) and diagnoses (based on ICD-10 codes) were available from 1 year before the index operation to the end of the study. To evaluate the diagnoses, we used the 10^th^ revision of the International Statistical Classification of Diseases and Related Health Problems (ICD-10 from 2019), which is available at [**https://icd.who.int/browse10/2019/en**](https://icd.who.int/browse10/2019/en)

The corresponding German version is available at [**https://www.dimdi.de/static/de/klassifikationen/icd/icd-10-who/kode-suche/htmlamtl2019/**](https://www.dimdi.de/static/de/klassifikationen/icd/icd-10-who/kode-suche/htmlamtl2019/).

The following table provides the MEL codes of surgeries used for the inclusion of patients in the AUTHEARTVISIT Study.

| **MEL Code** | **Description** |
| --- | --- |
| DB020 | Percutaneous implantation of a pulmonary valve |
| DB025 | Aortic valve replacement – catheter directed, transapical, TAVR |
| DB026 | Aortic valve replacement – catheter directed, transvalvular, TAVR |
| DB030 | Reconstruction of the aortic valve |
| DB040 | Reconstruction of the mitral valve |
| DB050 | Reconstruction of the tricuspid valve |
| DB055 | Reconstruction of the pulmonary valve |
| DB060 | Replacement of aortic valve with pulmonary autograft |
| DB070 | Replacement of aortic valve with stentless valve |
| DB080 | Replacement of aortic valve with stented valve |
| DB082 | Replacement of aortic valve with artificial mechanical valve |
| DB090 | Replacement of mitral valve with stentless valve |
| DB100 | Replacement of mitral valve with stented valve |
| DB102 | Replacement of mitral valve with artificial mechanical valve |
| DB110 | Replacement of tricuspid valve with stentless valve |
| DB120 | Replacement of tricuspid valve with stented valve |
| DB122 | Replacement of tricuspid valve with artificial mechanical valve |
| DB130 | Replacement of pulmonary valve with stentless valve |
| DB140 | Replacement of pulmonary valve with stented valve |
| DB142 | Replacement of pulmonary valve with artificial mechanical valve |
| DB021 | Aortic valve replacement – percutaneous, interventional, TAVR |
| XN010 | Aortic valve replacement – percutaneous, interventional, TAVR |

**Supplementary Table 1:** MEL codes used for inclusion and exclusion criteria as well as for definitions of outcomes and confounders.

2.) **Inclusion and Exclusion Criteria**

For the presented analyses of the AUTHEARTVISIT Study, clinical and operative data were obtained for all patients registered in the Austrian Health Care System who underwent surgical aortic valve replacement (AVR) using a mechanical prosthesis (sM-AVR, MEL code DB082) or biological prosthesis (sB-AVR, MEL codes DB060, DB070, and DB080) in Austria in the years 2010 to 2020 and were aged between 50 and 65 years. For a description of the MEL codes, see Supplementary Table 1.

Patients receiving transcatheter aortic valve replacement (TAVR; MEL codes DB025, DB026, DB021, or XN010, see Supplementary Table 1) as the index operation were not included in the sample. Patients aged <50 years and >65 years were also excluded from the data.

Patients with concomitant heart surgery were excluded from the data, i.e. patients with at least one valve surgery in addition to the index operation (sM-AVR or sB-AVR) on the date of the index operation (MEL codes listed in Supplementary Table 1).

Furthermore, patients receiving a coronary artery stent (MEL code DD050 “implantation of a stent in the coronary artery” or DD060 “implantation of a drug eluting stent in a coronary artery”) within 4 months prior to the AVR were excluded from this analysis. We selected a period of 4 months prior to the surgery for the exclusion of patients with percutaneous coronary intervention to guarantee the selection of patients with pure AVR procedures.

3.) **Definition of Outcomes**

For each patient, billing information (based on MEL codes) and diagnoses (based on ICD-10 codes) were available from 1 year before the index operation to the end of the study. Death dates were available until the end of the study. To evaluate the different outcomes for each patient, data were scanned for the corresponding codes from the index operation to the end of the study as shown in the following tables.

| **Outcome** | **Definition** |
| --- | --- |
| Death | All-cause death based on death data |
| Reoperation | Based on billing information (MEL code): first event after index operation with MEL code defined as in Supplementary Table 1 |
| Death or reoperation | Combined endpoint: first event after index operation with MEL code defined as in Supplementary Table 1 or death based on death date |
| Myocardial infarction | Based on ICD codes: first event after index operation with ICD code defined as in Supplementary Table 3 |
| Heart failure | Based on ICD codes: first event after index operation with ICD code defined as in Supplementary Table 3 |
| Embolic stroke or intracerebral hemorrhage (ICH) | Based on ICD codes: first event after index operation with ICD code defined as in Supplementary Table 3 |
| Bleeding other than ICH | Based on ICD codes: first event after index operation with ICD code defined as in Supplementary Table 3 |
| MACE | Combined endpoint: first event after index operation with ICD codes for myocardial infarction, heart failure, embolic stroke or ICH, reoperation, or death |
| ICH | Based on ICD codes: first event after index operation with ICD code defined as in Supplementary Table 3 |
| Death after reoperation | Time from reoperation defined as in Supplementary Table 1 to death date |

**Supplementary Table 2:** Definitions of outcomes

| **Outcome** | **ICD-10 Codes** |
| --- | --- |
| Myocardial infarction | I21.0, I21.1, I21.2, I21.3, I21.4, I21.9 |
| Heart failure | I11.0, I13.0, I13.2, I50.0, I50.1, I50.9, I50.11, I50.12, I50.13, I50.14, I50.19 |
| Embolic stroke or intracerebral hemorrhage (ICH) | I63.0, I63.1, I63.2, I63.3, I63.4, I63.5, I63.6, I63.8, I63.9, G45.9, G45.0, G45.1, G45.2, G45.3, G45.4, G45.8, I61.0, I61.1, I61.2, I61.3, I61.4, I61.5, I61.6, I61.8, I61.9, I64 |
| Bleeding other than ICH | I60.0, I60.1, I60.2, I60.3, I60.4, I60.5, I60.6, I60.7, I60.8, I60.9, I85.0, I98.2, I98.3, K25.0, K25.1, K25.2, K25.3, K25.4, K25.5, K25.6, K25.7, K25.9, K26.0, K26.1, K26.2, K26.3, K26.4, K26.5, K26.6, K26.7, K26.9, K27.0, K27.1, K27.2, K27.3, K27.4, K27.5, K27.6, K27.7, K27.9, K28.0, K28.1, K28.2, K28.3, K28.4, K28.5, K28.6, K28.7, K28.9, K29.0, K29.1, K29.2, K29.3, K29.4, K29.5, K29.6, K92.2, N42.1, R04.1, R04.8, R04.9, R58, S06.4, T81.0 |
| ICH | I61.0, I61.1, I61.2, I61.3, I61.4, I61.5, I61.6, I61.8, I61.9 |

**Supplementary Table 3:** Definitions of outcomes based on ICD-10 codes

4.) **Definitions of Confounders/Comorbidities**

The index operation group was defined using billing information based on MEL codes as in Supplementary Table 4.

| **MEL Code** | **Description** | **Group** |
| --- | --- | --- |
| DB060 | Replacement of aortic valve with pulmonary autograft | sB-AVR |
| DB070 | Replacement of aortic valve with stentless valve | sB-AVR |
| DB080 | Replacement of aortic valve with stented valve | sB-AVR |
| DB082 | Replacement of aortic valve with artificial mechanical valve | sM-AVR |

**Supplementary Table 4:** Coding for prosthesis variable. sB-AVR = bioprosthesis; sM-AVR = mechanical aortic valve replacement

Comorbidities were defined using ICD-10 codes available for each patient up to 1 year before the index operation. Data available 1 year before the index operation were scanned for each patient based on the following ICD-10 codes, categorized for different comorbidities (Supplementary Table 5). If at least once during the year prior to the index operation an ICD-10 code for a comorbidity was observed as the main or secondary diagnosis, the patient was assumed to suffer from this comorbidity.

| Comorbidity | ICD Codes |
| --- | --- |
| Diabetes mellitus | E10.0, E10.1, E10.2, E10.3, E10.4, E10.5, E10.6, E10.7, E10.8, E10.9, E11.0, E11.1, E11.2, E11.3, E11.4, E11.5, E11.6, E11.7, E11.8, E11.9, E12.0, E12.1, E12.2, E12.3, E12.4, E12.5, E12.6, E12.7, E12.8, E12.9, E13.0, E13.1, E13.2, E13.3, E13.4, E13.5, E13.6, E13.7, E13.8, E13.9, E14.0, E14.1, E14.2, E14.3, E14.4, E14.5, E14.6, E14.7, E14.8, E14.9 |
| Adiposity | E65, E66.0, E66.1, E66.2, E66.8, E66.9 |
| Hyperlipidemia | E78.0, E78.1, E78.2, E78.3, E78.4, E78.5, E78.6, E78.8, E78.9 |
| Hyperuricemia/gout | E79.0, E79.8, M10.0, M10.00, M10.01, M10.02, M10.03, M10.04, M10.05, M10.06, M10.07, M10.08, M10.09 |
| Valvular, rhythmological, and other cardiomyopathies (CMPs) | I01.0, I01.1, I01.2, I01.8, I01.9, I02.0, I02.9, I05.0, I05.1, I05.2, I05.8, I05.9, I06.0, I06.1, I06.2, I06.8, I06.9, I07.0, I07.1, I07.2, I07.8, I07.9, I08.0, I08.1, I08.2, I08.3, I08.8, I08.9, I09.0, I09.1, I09.2, I09.8, I09.9, I10, I11.0, I11.9, I12.0, I12.9, I13.0, I13.1, I13.2, I13.9, I15.0, I15.1, I15.2, I15.8, I15.9, I26.0, I26.9, I27.0, I27.1, I27.2, I27.8, I27.9, I28.0, I28.1, I28.8, I28.9, I30.0, I30.1, I30.8, I30.9, I31.0, I31.1, I31.2, I31.3, I31.8, I31.9, I32.0, I32.1, I32.8, I33.0, I33.9, I34.0, I34.1, I34.2, I34.8, I34.9, I35.0, I35.1, I35.2, I35.8, I35.9, I36.0, I36.1, I36.2, I36.8, I36.9, I37.0, I37.1, I37.2, I37.8, I37.9, I38, I39.0, I39.1, I39.2, I39.3, I39.4, I39.8, I40.0, I40.1, I40.8, I40.9, I41.0, I41.1, I41.2, I41.8, I42.0, I42.1, I42.2, I42.3, I42.4, I42.5, I42.6, I42.7, I42.8, I42.9, I43.0, I43.1, I43.2, I43.8, I44.0, I44.1, I44.2, I44.3, I44.4, I44.5, I44.6, I44.7, I45.0, I45.1, I45.2, I45.3, I45.4, I45.5, I45.6, I45.8, I45.9, I46.0, I46.1, I46.9, I470., I47.1, I47.2, I47.9, I48.0, I48.1, I48.2, I48.3, I48.4, I48.9, I49.0, I49.1, I49.2, I49.3, I49.4, I49.5, I49.8, I49.9, I50.0, I50.11, I50.12, I50.13, I50.14, I50.19, I50.9, I51.0, I51.1, I51.2, I51.3, I51.4, I51.5, I51.6, I51.7, I51.8, I51.9, I52.0, I52.1, I52.8, Q20.0, Q20.1, Q20.2, Q20.3, Q20.4, Q20.5, Q20.6, Q20.8, Q20.9, Q21.0, Q21.1, Q21.2, Q21.3, Q21.4, Q21.8, Q21.9, Q22.0, Q22.1, Q22.2, Q22.3, Q22.4, Q22.5, Q22.6, Q22.8, Q22.9, Q23.0, Q23.1, Q23.2, Q23.3, Q23.4, Q23.8, Q23.9, Q24.0, Q24.1, Q24.2, Q24.3, Q24.4, Q24.5, Q24.6, Q24.8, Q24.9, Q25.0, Q25.1, Q25.2, Q25.3, Q25.4, Q25.5, Q25.6, Q25.7, Q25.8, Q25.9 |
| Atherosclerosis | I69.8, I70.0, I70.1, I70.2, I70.8, I70.9 |
| Pulmonary disease | J43.1, J43.2, J43.8, J43.9, J44.00, J44.01, J44.02, J44.03, J44.09, J44.10, J44.11, J44.12, J44.13, J44.19, J44.80, J44.81, J44.82, J44.83, J44.89, J44.90, J44.91, J44.92, J44.93, J44.99, J45.0, J45.1, J45.8, J45.9 |
| Kidney disease | N00,0, N00.1, N00.2, N00.3, N00.4, N00.5, N00.6, N00.7, N00.8, N00.9, N01.0, N01.1, N01.2, N01.3, N01.4, N01.5, N01.6, N01.7, N01.8, N01.9, N02.0, N02.1, N02.2, N02.3, N02.4, N02.5, N02.6, N02.7, N02.8, N02.9, N03.0, N03.1, N03.2, N03.3, N03.4, N03.5, N03.6, N03.7, N03.8, N03.9, N04.0, N04.1, N04.2, N04.3, N04.4, N04.5, N04.6, N04.7, N04.8, N04.9, N05.0, N05.1, N05.2, N05.3, N05.4, N05.5, N05.6, N05.7, N05.8, N05.9, N06.0, N06.1, N06.2, N06.3, N06.4, N06.5, N06.6, N06.7, N06.8, N06.9, N07.0, N07.1, N07.2, N07.3, N07.4, N07.5, N07.6, N07.7, N07.8, N07.9, N08.0, N08.1, N08.2, N08.3, N08.4, N08.5, N08.8, N10, N11.0, N11.1, N11.8, N11.9, N12, N13.0, N13.1, N13.2, N13.3, N13.4, N13.5, N13.6, N13.7, N13.8, N13.9, N14.0, N14.1, N14.2, N14.3, N14.4, N15.0, N15.1, N15.8, N15.9, N16.0, N16.1, N16.2, N16.3, N16.4, N16.5, N16.8, N17.0, N17.1, N17.2, N17.8, N17.9, N18.1, N18.2, N18.3, N18.4, N18.5, N18.9, N19, N20.0 |
| Ischemic cardiomyopathies (CMP) | I20.0, I20.1, I20.8, I20.9, I21.0, I21.1, I21.2, I21.3, I21.4, I21.9, I22.0, I22.1, I22.8, I22.9, I23.0, I23.1, I23.2, I23.3, I23.4, I23.5, I23.6, I23.8, I24.0, I24.1, I24.8, I24.9, I25.0, I25.1, I25.2, I25.3, I25.4, I25.5, I25.6, I25.8, I25.9 |
| Malignant diseases | C00.0, C00.1, C00.2, C00.3, C00.4, C00.5, C00.6, C00.8, C00.9, C01, C02.0, C02.1, C02.2, C02.3, C02.4, C02.8, C02.9, C03.0, C03.1, C03.9, C04.0, C04.1, C04.8, C04.9, C05.0, C05.1, C05.2, C05.8, C05.9, C06.0, C06.1, C06.2, C06.8, C06.9, C07 , C08.0, C08.1, C08.8, C08.9, C09.0, C09.1, C09.8, C09.9, C10.0, C10.1, C102., C10.3, C10.4, C10.8, C10.9, C11.0, C11.1, C11.2, C11.3, C11.8, C11.9, C12, C13.0, C13.1, C13.2, C13.8, C13.9, C14.0, C14.2, C14.8, C15.0, C15.1, C15.2, C15.3, C15.4, C15.5, C15.8, C15.9, C16.0, C16.1, C16.2, C16.3, C16.4, C16.5, C16.6, C16.8, C16.9, C17.0, C17.1, C17.2, C17.3, C17.8, C17.9, C18.0, C180.1,  C18.02, C180.3, C18.04, C18.1, C18.11, C18.12, C18.13, C18.14, C182., C18.21, C18.22, C18.23, C18.24, C18.3, C18.31, C18.32, C18.33, C18.34, C18.4, C18.41, C18.42, C18.43, C18.44, C18.5, C18.51, C18.52, C18.53, C18.54, C18.6, C18.61, C18.62, C18.63, C18.64, C18.7, C18.71, C18.72, C18.73, C18.74, C18.8, C18.81, C18.82, C18.83, C18.84, C18.9, C18.91, C18.92, C18.93, C18.94, C19, C19.1, C19.2, C19.3, C19.4, C20, C20.1, C20.2, C20.3, C20.4, C21.0, C21.1, C21.2, C21.8, C22.0, C22.1, C22.2, C22.3, C22.4, C22.7, C22.9, C23, C24.0, C24.1, C24.8, C24.9, C25.0, C25.1, C25.2, C25.3, C25.4, C25.7, C25.8, C25.9, C26.0, C26.1, C26.8, C26.9, C30.0, C30.1, C31.0, C31.1, C31.2, C31.3, C31.8, C31.9, C32.0, C32.1, C32.2, C32.3, C32.8, C32.9, C33, C34.0, C34.1, C34.2, C34.3, C34.8, C34.9, C37, C38.0, C38.1, C38.2, C38.3, C38.4, C38.8, C39.0, C39.8, C39.9, C40.0, C40.1, C40.2, C40.3, C40.8, C40.9, C41.0, C41.1, C41.2, C41.3, C41.4, C41.8, C41.9, C43.0, C43.1, C43.2, C43.3, C43.4, C43.5, C43.6, C43.7, C43.8, C43.9, C44.0, C44.1, C44.2, C44.3, C44.4, C44.5, C44.6, C44.7, C44.8, C44.9, C45.0, C45.1, C45.2, C45.7, C45.9, C46.0, C46.1, C46.2, C46.3, C46.7, C46.8, C46.9, C47.0, C47.1, C47.2, C47.3, C47.4, C47.5, C47.6, C47.8, C47.9, C48.0, C48.1, C48.2, C48.8, C49.0, C49.1, C49.2, C49.3, C49.4, C49.5, C49.6, C49.8, C49.9, C50.0, C50.1, C50.2, C50.3, C50.4, C50.5, C50.6, C50.8, C50.9, C51.0, C51.1, C51.2, C51.8, C51.9, C52, C53.0, C53.1, C53.8, C53.9, C54.0, C54.1, C54.2, C54.3, C54.8, C54.9, C55, C56, C57.0, C57.1, C57.2, C57.3, C57.4, C57.7, C57.8, C57.9, C58, C60.0, C60.1, C60.2, C60.8, C609., C61, C62.0, C62.1, C62.9, C63.0, C63.1, C63.2, C63.7, C63.8, C63.9, C64, C65, C66, C67.0, C67.1, C67.2, C67.3, C67.4, C67.5, C67.6, C67.7, C67.8, C67.9, C68.0, C68.1, C68.8, C68.9, C69.0, C69.1, C69.2, C69.3, C69.4, C69.5, C69.6, C69.8, C69.9, C70.0, C70.1, C70.9, C71.0, C71.1, C71.2, C71.3, C71.4, C71.5, C71.6, C71.7, C71.8, C71.9, C72.0, C72.1, C72.2, C72.3, C72.4, C72.5, C72.8, C72.9, C73, C740., C74.1, C749., C75.0, C75.1, C75.2, C75.3, C75.4, C75.5, C75.8, C75.9, C76.0, C76.1, C76.2, C76.3, C76.4, C76.5, C76.7, C76.8, C77.0, C77.1, C77.2, C77.3, C77.4, C77.5, C77.8, C77.9, C78.0, C78.1, C78.2, C78.3, C78.4, C78.5, C78.6, C78.7, C78.8, C79.0, C79.1, C79.2, C79.3, C79.4, C79.5, C79.6, C79.7, C79.8, C79.9, C80.0, C80.9, C81.0, C81.1, C81.2, C81.3, C81.4, C81.7, C81.9, C82.0, C82.1, C82.2, C82.3, C82.4, C82.5, C82.6, C82.7, C82.9, C83.0, C83.1, C83.3, C83.5, C83.7, C83.8, C83.9, C840., C84.1, C84.4, C845., C84.6, C84.7, C84.8, C84.9, C85.1, C85.2, C85.7, C85.9, C86.0, C86.1, C86.2, C86.3, C86.4, C86.5, C86.6, C88.0, C88.2, C88.3, C88.4, C887., C88.9, C90.0, C90.1, C90.2, C90.3, C91.0, C91.1, C91.3, C91.4, C91.5, C91.6, C91.7, C91.8, C91.9, C92.0, C92.1, C92.2, C92.3, C92.4, C92.5, C92.6, C92.7, C92.8, C92.9, C93.0, C93.1, C93.3, C93.7, C93.9, C94.0, C94.2, C94.3, C94.4, C94.6, C94.7, C95.0, C95.1, C95.7, C95.9, C96.0, C96.2, C96.4, C96.5, C96.6, C96.7, C96.8, C96.9, C97 |
| Stroke before OP | ICD-Codes as in Table 1 |
| Heart failure before OP | ICD-Codes as in Table 1 |
| Heart attack before OP | ICD-Codes as in Table 1 |

**Supplementary Table 5:** ICD-10 codes for comorbidities

5.) **Medication before Index Surgery**

| Medication | sM-AVR (n = 1018) | sB-AVR (n = 2743) | P-value |
| --- | --- | --- | --- |
| Therapies for hepatobiliary diseases | 4 (0.39%) | 8 (0.29%) | 0.870 |
| Intestinal anti-infectives | 9 (0.88%) | 36 (1.31%) | 0.366 |
| Intestinal anti-inflammatory drugs (steroids) | 5 (0.49%) | 8 (0.29%) | 0.539 |
| Insulins | 37 (3.63%) | 160 (5.83%) | 0.009 |
| Non-insulin antidiabetics (e.g., SLGT2i and GLP1-RA) | 111 (10.9%) | 421 (15.35%) | <0.001 |
| Vitamin D and analogues | 27 (2.65%) | 124 (4.52%) | 0.012 |
| Calcium, potassium, magnesium salts | 58 (5.7%) | 218 (7.95%) | 0.023 |
| Vitamin K antagonists | 76 (7.47%) | 121 (4.41%) | <0.001 |
| Heparins (UFH and LMWH) | 160 (15.72%) | 372 (13.56%) | 0.103 |
| Platelet aggregation inhibitors | 110 (10.81%) | 410 (14.95%) | 0.001 |
| Direct thrombin inhibitors and Fxa inhibitors (anticoagulants) | 51 (5.01%) | 129 (4.7%) | 0.760 |
| Iron substitution | 16 (1.57%) | 68 (2.48%) | 0.121 |
| Anti-anemic drugs (ESA, HIFi) | 5 (0.49%) | 37 (1.35%) | 0.04 |
| Glycosides | 12 (1.18%) | 30 (1.09%) | 0.963 |
| Antiarrhythmic drugs (excluding digitalis glycosides) | 21 (2.06%) | 83 (3.03%) | 0.137 |
| Adrenergic and dopaminergic stimulants (including PDE) | 6 (0.59%) | 15 (0.55%) | 1 |
| Vasodilators | 58 (5.7%) | 230 (8.38%) | 0.007 |
| Antihypertensives (excluding BB, MRA, loop diuretics, RAASi) | 170 (16.7%) | 553 (20.16%) | 0.019 |
| New therapeutics for PHA | 0 (0%) | 2 (0.07%) | 0.948 |
| Loop diuretics | 67 (6.58%) | 216 (7.87%) | 0.205 |
| MRA | 85 (8.35%) | 236 (8.6%) | 0.856 |
| Beta-blockers | 296 (29.08%) | 814 (29.68%) | 0.751 |
| RAAS inhibitors | 446 (43.81%) | 1240 (45.21%) | 0.467 |
| Fat-lowering agents (statins, fibrates, and others) | 459 (45.09%) | 1350 (49.22%) | 0.027 |
| Topical antibiotics | 17 (1.67%) | 57 (2.08%) | 0.504 |
| Dermal steroids | 48 (4.72%) | 181 (6.6%) | 0.039 |
| Parathyroid antagonists | 5 (0.49%) | 25 (0.91%) | 0.28 |
| Systemic antibiotics | 436 (42.83%) | 1245 (45.39%) | 0.172 |
| Systemic antiviral drugs | 24 (2.36%) | 63 (2.3%) | 1 |
| Chemotherapies, including immunotherapies | 3 (0.29%) | 30 (1.09%) | 0.033 |
| Immunosuppressants | 30 (2.95%) | 81 (2.95%) | 1 |
| NSAIDs | 283 (27.8%) | 855 (31.17%) | 0.05 |
| Gout medication | 44 (4.32%) | 149 (5.43%) | 0.198 |
| Biophosphonates | 12 (1.18%) | 49 (1.79%) | 0.244 |
| Non-NSAID analgesics, including opioids | 132 (12.97%) | 341 (12.43%) | 0.701 |
| Inhalants against COPD | 174 (17.09%) | 598 (21.8%) | 0.002 |
| Xanthines and leukotriene antagonists | 15 (1.47%) | 47 (1.71%) | 0.712 |
| Direct thrombin inhibitors and Fxa inhibitors (anticoagulants) or vitamin K antagonists | 126 (12.38%) | 244 (8.9%) | 0.002 |

**Supplementary Table 6:** Prescribed medications the year prior to the index surgery

6.) **Primary Outcome: All-Cause Death**

|  | Patients aged 50 – 65 years | | | | Patients aged 50 – 60 years | | | |
| --- | --- | --- | --- | --- | --- | --- | --- | --- |
|  | All data | | After PSM | | All data | | After PSM | |
| Variables | HR (95% CI) | P-value | HR (95% CI) | P-value | HR (95% CI) | P-value | HR (95% CI) | P-value |
| Original Model | | | | | | | | |
| Heart valve (sM-AVR) | 1.352  (1.109 - 1.649) | 0.003 | 1.400  (1.103 - 1.778) | 0.006 | 1.601  (1.235 - 2.074) | <0.001 | 1.645  (1.209 - 2.238) | 0.002 |
| Age | 1.036  (1.016 - 1.057) | <0.001 | 1.067  (1.036 - 1.099) | <0.001 | 1.054  (1.013 - 1.097) | 0.009 | 1.070  (1.014 - 1.128) | 0.013 |
| Sex (M) | 1.011  (0.849 - 1.205) | 0.900 | 0.804  (0.606 - 1.068) | 0.133 | 1.122  (0.862 - 1.459) | 0.392 | 0.978  (0.687 - 1.392) | 0.902 |
| Heart failure (Yes) | 1.643  (1.328 - 2.033) | <0.001 | 1.469  (1.022 - 2.111) | 0.038 | 1.451  (1.017 - 2.07) | 0.040 | 1.217  (0.691 - 2.144) | 0.497 |
| Myocardial infarction (Yes) | 1.068  (0.718 - 1.587) | 0.745 | 0.993  (0.496 - 1.988) | 0.983 | 0.815  (0.395 - 1.682) | 0.581 | 0.675  (0.164 - 2.779) | 0.586 |
| Embolic stroke or ICH (Yes) | 1.699  (1.016 - 2.841) | 0.043 | 2.909  (1.264 - 6.698) | 0.012 | 1.709  (0.874 - 3.344) | 0.117 | 2.902  (0.902 - 9.339) | 0.074 |
| Diabetes mellitus (Yes) | 1.640  (1.35 - 1.992) | <0.001 | 1.407  (1.014 - 1.952) | 0.041 | 1.703  (1.246 - 2.328) | 0.001 | 1.675  (1.069 - 2.625) | 0.024 |
| Adiposity (Yes) | 1.430  (1.123 - 1.821) | 0.004 | 1.532  (1.037 - 2.263) | 0.032 | 1.458  (1.01 - 2.105) | 0.044 | 1.131  (0.634 - 2.017) | 0.678 |
| Hyperlipidemia (Yes) | 0.674  (0.551 - 0.824) | <0.001 | 0.720  (0.527 - 0.985) | 0.040 | 0.820  (0.609 - 1.102) | 0.188 | 0.952  (0.637 - 1.423) | 0.811 |
| Hyperuricemia/gout (Yes) | 1.271  (0.871 - 1.855) | 0.213 | 1.320  (0.792 - 2.201) | 0.287 | 1.471  (0.783 - 2.765) | 0.23 | 1.129  (0.468 - 2.726) | 0.787 |
| Valvular, rhythmological, and other CMPs (Yes) | 0.684  (0.552 - 0.848) | 0.001 | 0.800  (0.580 - 1.104) | 0.174 | 0.608  (0.449 - 0.825) | 0.001 | 0.591  (0.407 - 0.86) | 0.006 |
| Ischemic CMP (Yes) | 1.154  (0.977 - 1.362) | 0.092 | 1.138  (0.877 - 1.477) | 0.332 | 1.171  (0.912 - 1.504) | 0.215 | 1.052  (0.758 - 1.461) | 0.761 |
| Atherosclerosis (Yes) | 1.452  (1.063 - 1.984) | 0.019 | 1.266  (0.700 - 2.288) | 0.435 | 1.577  (0.926 - 2.686) | 0.093 | 2.120  (1.068 - 4.206) | 0.032 |
| Pulmonary diseases (Yes) | 2.014  (1.405 - 2.889) | <0.001 | 1.563  (0.819 - 2.983) | 0.176 | 2.136  (1.202 - 3.795) | 0.010 | 2.807  (1.328 - 5.935) | 0.007 |
| Kidney diseases (Yes) | 2.330  (1.851 - 2.932) | <0.001 | 2.541  (1.765 - 3.659) | <0.001 | 2.371  (1.658 - 3.391) | <0.001 | 2.611  (1.575 - 4.329) | <0.001 |
| Malignant diseases (Yes) | 1.328  (0.928 - 1.9) | 0.121 | 0.546  (0.199 - 1.494) | 0.238 | 1.811  (1.079 - 3.039) | 0.025 | 2.859  (1.026 - 7.966) | 0.045 |
| Interaction Terms | | | | | | | | |
| Heart valve (sM-AVR) × Age | 0.965  (0.922 - 1.009) | 0.118 | 0.979  (0.923 - 1.037) | 0.466 | 1.017  (0.933 - 1.108) | 0.703 | 1.046  (0.941 - 1.162) | 0.403 |
| Heart valve (sM-AVR) × Sex (M) | 1.105  (0.718 - 1.702) | 0.649 | 0.974  (0.557 - 1.701) | 0.926 | 1.164  (0.651 - 2.080) | 0.609 | 0.854  (0.423 - 1.725) | 0.660 |

**Supplementary Table 7:** Hazard ratios (HRs) and corresponding 95% confidence intervals (CIs) from multivariable Cox regression accounting for all listed confounders for all-cause mortality in all patients and the subgroup of patients aged 50 - 60 years before and after propensity score matching (PSM)

7.) **Secondary Outcomes**

**7.1. MACEs**

|  | Patients aged 50 – 65 years | | | | Patients aged 50 – 60 years | | | | |
| --- | --- | --- | --- | --- | --- | --- | --- | --- | --- |
|  | All data | | After PSM | | All data | | | After PSM | |
| Variables | HR (95% CI) | P-value | HR (95% CI) | P-value | HR (95% CI) | | P-value | HR (95% CI) | P-value |
| Original Model | | | | | | | | | |
| Heart valve (sM-AVR) | 1.182  (1.016 - 1.375) | 0.030 | 1.224  (1.020 - 1.469) | 0.029 | 1.340  (1.105 - 1.624) | 0.003 | | 1.370  (1.092 - 1.718) | 0.007 |
| Age | 1.029  (1.013 - 1.044) | <0.001 | 1.032  (1.008 - 1.055) | 0.007 | 1.027  (0.996 - 1.059) | 0.089 | | 1.031  (0.990 - 1.073) | 0.137 |
| Sex (M) | 1.008  (0.878 - 1.157) | 0.910 | 0.860  (0.694 - 1.066) | 0.168 | 1.101  (0.899 - 1.349) | 0.353 | | 1.003  (0.771 - 1.305) | 0.982 |
| Heart failure (Yes) | 2.079  (1.762 - 2.452) | <0.001 | 2.628  (2.037 - 3.391) | <0.001 | 2.047  (1.577 - 2.658) | <0.001 | | 2.269  (1.567 - 3.287) | <0.001 |
| Myocardial infarction (Yes) | 1.430  (1.061 - 1.926) | 0.019 | 1.528  (0.931 - 2.508) | 0.093 | 1.054  (0.629 - 1.767) | 0.840 | | 1.315  (0.573 - 3.015) | 0.518 |
| Embolic stroke or ICH (Yes) | 1.885 (1.283 - 2.768) | 0.001 | 2.061  (1.012 - 4.198) | 0.046 | 1.732  (1.010 - 2.97) | 0.046 | | 1.377  (0.432 - 4.394) | 0.589 |
| Diabetes mellitus (Yes) | 1.595  (1.366 - 1.862) | <0.001 | 1.400  (1.087 - 1.804) | 0.009 | 1.599  (1.252 - 2.042) | <0.001 | | 1.612  (1.150 - 2.26) | 0.006 |
| Adiposity (Yes) | 1.372  (1.128 - 1.668) | 0.002 | 1.381  (1.008 - 1.893) | 0.045 | 1.427  (1.068 - 1.908) | 0.016 | | 1.081  (0.693 - 1.686) | 0.731 |
| Hyperlipidemia (Yes) | 0.818  (0.703 - 0.953) | 0.010 | 0.883  (0.699 - 1.116) | 0.298 | 0.913  (0.73 - 1.142) | 0.424 | | 0.934  (0.694 - 1.256) | 0.650 |
| Hyperuricemia/gout (Yes) | 1.217  (0.896 - 1.653) | 0.209 | 1.099  (0.715 - 1.69) | 0.667 | 1.369  (0.841 - 2.230) | 0.207 | | 0.977  (0.486 - 1.964) | 0.949 |
| Valvular, rhythmological, and other CMPs (Yes) | 0.776  (0.653 - 0.921) | 0.004 | 0.858  (0.667 - 1.103) | 0.233 | 0.710  (0.559 - 0.903) | 0.005 | | 0.748  (0.558 - 1.002) | 0.051 |
| Ischemic CMP (Yes) | 0.982  (0.861 - 1.120) | 0.788 | 1.033  (0.846 - 1.262) | 0.747 | 1.091  (0.900 - 1.322) | 0.376 | | 0.986  (0.772 - 1.260) | 0.910 |
| Atherosclerosis (Yes) | 1.463  (1.139 - 1.880) | 0.003 | 1.435  (0.927 - 2.221) | 0.105 | 1.156  (0.740 - 1.805) | 0.525 | | 1.574  (0.891 - 2.78) | 0.118 |
| Pulmonary diseases (Yes) | 1.222  (0.884 - 1.688) | 0.225 | 1.042  (0.595 - 1.824) | 0.885 | 1.348  (0.833 - 2.182) | 0.223 | | 1.645 (0.89 - 3.039) | 0.112 |
| Kidney diseases (Yes) | 1.843  (1.516 - 2.240) | <0.001 | 1.899  (1.399 - 2.577) | <0.001 | 2.008  (1.491 - 2.705) | <0.001 | | 2.205  (1.469 - 3.310) | <0.001 |
| Malignant diseases (Yes) | 1.020  (0.738 - 1.408) | 0.905 | 0.436  (0.179 - 1.061) | 0.067 | 0.981  (0.590 - 1.629) | 0.939 | | 1.155  (0.424 - 3.149) | 0.778 |
| Interaction Terms | | | | | | | | | |
| Heart valve (sM-AVR) × Age | 0.965  (0.933 - 0.999) | 0.042 | 0.959  (0.918 - 1.002) | 0.063 | 0.969  (0.909 - 1.032) | 0.329 | | 0.972  (0.899 - 1.051) | 0.480 |
| Heart valve (sM-AVR) × Sex (M) | 1.067  (0.770 - 1.478) | 0.696 | 0.991  (0.650 - 1.511) | 0.967 | 1.078  (0.700 - 1.659) | 0.734 | | 0.775  (0.461 - 1.304) | 0.337 |

**Supplementary Table 8:** Hazard ratios (HRs) and corresponding 95% confidence intervals (CIs) from multivariable Cox regression accounting for all listed confounders for MACEs in all patients and the subgroup of patients aged 50 – 60 years before and after propensity score matching (PSM)

**Figure S1:** Kaplan-Meier curves and 95% confidence intervals for MACEs before (A,C) and after (B,D) PSM for all patients aged 50 – 65 years (A,B) and the subgroup of patients aged 50 – 60 years (C,D)

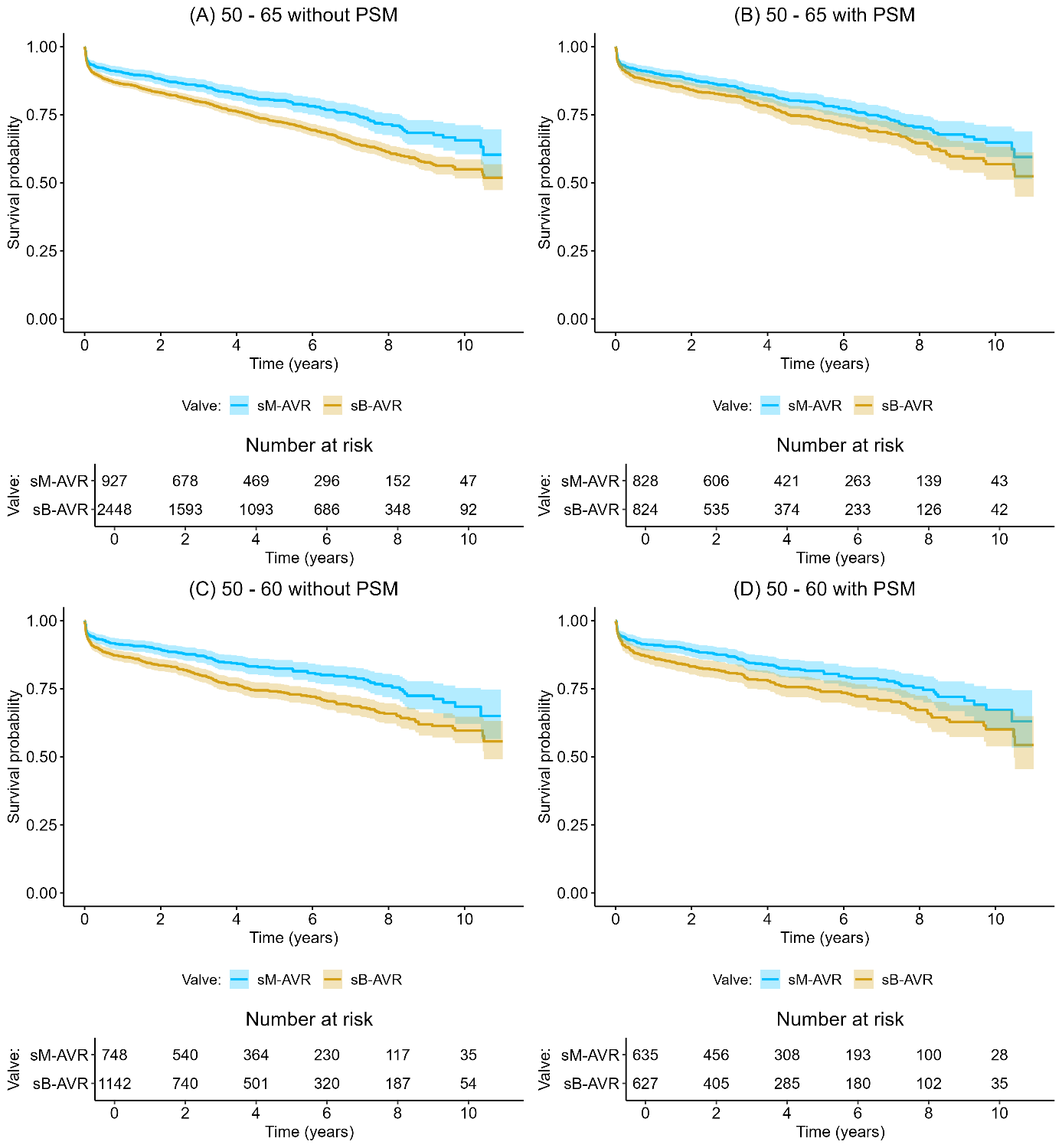

**7.2. Death or reoperation**

|  | Patients aged 50 – 65 years | | | | Patients aged 50 – 60 years | | | |
| --- | --- | --- | --- | --- | --- | --- | --- | --- |
|  | All data | | After PSM | | All data | | After PSM | |
| Variables | HR (95% CI) | P-value | HR (95% CI) | P-value | HR (95% CI) | P-value | HR (95% CI) | P-value |
| Original Model | | | | | | | | |
| Heart valve (sM-AVR) | 1.550  (1.26 - 1.908) | <0.001 | 1.566  (1.223 - 2.005) | <0.001 | 1.732  (1.325 - 2.264) | <0.001 | 1.885  (1.373 - 2.587) | <0.001 |
| Age | 1.034  (1.014 - 1.054) | 0.001 | 1.056  (1.024 - 1.088) | 0.001 | 1.049  (1.007 - 1.092) | 0.022 | 1.069  (1.013 - 1.128) | 0.015 |
| Sex (M) | 1.037  (0.870 - 1.237) | 0.682 | 0.885  (0.665 - 1.178) | 0.402 | 1.156  (0.886 - 1.507) | 0.286 | 1.013  (0.709 - 1.448) | 0.943 |
| Heart failure (Yes) | 1.583  (1.271 - 1.971) | <0.001 | 1.562  (1.082 - 2.256) | 0.017 | 1.548  (1.082 - 2.216) | 0.017 | 1.327  (0.753 - 2.337) | 0.328 |
| Myocardial infarction (Yes) | 1.092  (0.729 - 1.636) | 0.671 | 1.128  (0.561 - 2.265) | 0.736 | 0.997  (0.500 - 1.985) | 0.992 | 1.070  (0.331 - 3.456) | 0.910 |
| Embolic stroke or ICH (Yes) | 1.938  (1.194 - 3.145) | 0.007 | 1.806  (0.657 - 4.965) | 0.252 | 1.449  (0.712 - 2.948) | 0.306 | 1.713  (0.414 - 7.089) | 0.458 |
| Diabetes mellitus (Yes) | 1.593  (1.305 - 1.945) | <0.001 | 1.479  (1.052 - 2.078) | 0.024 | 1.570  (1.135 - 2.171) | 0.006 | 1.715  (1.086 - 2.708) | 0.021 |
| Adiposity (Yes) | 1.277  (0.989 - 1.648) | 0.061 | 1.391  (0.915 - 2.115) | 0.122 | 1.234  (0.832 - 1.83) | 0.296 | 0.709  (0.358 - 1.406) | 0.325 |
| Hyperlipidemia (Yes) | 0.674  (0.548 - 0.828) | <0.001 | 0.693  (0.498 - 0.963) | 0.029 | 0.798  (0.587 - 1.084) | 0.149 | 0.979  (0.653 - 1.467) | 0.918 |
| Hyperuricemia/gout (Yes) | 1.071  (0.705 - 1.629) | 0.748 | 1.068  (0.593 - 1.926) | 0.826 | 1.282  (0.645 - 2.549) | 0.478 | 1.119  (0.436 - 2.871) | 0.816 |
| Valvular, rhythmological, and other CMPs (Yes) | 0.708  (0.571 - 0.878) | 0.002 | 0.831  (0.599 - 1.154) | 0.269 | 0.654  (0.481 - 0.889) | 0.007 | 0.648  (0.443 - 0.949) | 0.026 |
| Ischemic CMP (Yes) | 1.098  (0.927 - 1.300) | 0.279 | 1.067  (0.815 - 1.398) | 0.635 | 1.131  (0.876 - 1.461) | 0.344 | 1.019  (0.731 - 1.422) | 0.910 |
| Atherosclerosis (Yes) | 1.443  (1.045 - 1.992) | 0.026 | 1.422  (0.785 - 2.577) | 0.245 | 1.706  (1.000 - 2.911) | 0.050 | 2.456  (1.273 - 4.738) | 0.007 |
| Pulmonary diseases (Yes) | 1.774  (1.183 - 2.661) | 0.006 | 0.686  (0.253 - 1.859) | 0.459 | 1.938  (1.042 - 3.602) | 0.037 | 2.161  (0.931 - 5.017) | 0.073 |
| Kidney diseases (Yes) | 2.172  (1.707 - 2.763) | <0.001 | 2.241  (1.523 - 3.298) | <0.001 | 1.994  (1.356 - 2.933) | <0.001 | 2.090  (1.197 - 3.651) | 0.010 |
| Malignant diseases (Yes) | 1.210  (0.832 - 1.758) | 0.319 | 0.612  (0.224 - 1.669) | 0.337 | 1.683  (0.986 - 2.872) | 0.056 | 3.111  (1.119 - 8.648) | 0.030 |
| Interaction Terms | | | | | | | | |
| Heart valve (sM-AVR) × Age | 0.970  (0.925 - 1.017) | 0.204 | 0.985  (0.927 - 1.045) | 0.610 | 0.993  (0.908 - 1.085) | 0.869 | 0.997  (0.893 - 1.112) | 0.956 |
| Heart valve (sM-AVR) × Sex (M) | 1.286  (0.811 - 2.038) | 0.285 | 1.299  (0.734 - 2.301) | 0.369 | 1.309  (0.714 - 2.399) | 0.384 | 1.134  (0.546 - 2.354) | 0.737 |

**Supplementary Table 9:** Hazard ratios (HRs) and corresponding 95% confidence intervals (CIs) from multivariable Cox regression accounting for all listed confounders for death or reoperation in all patients and the subgroup of patients aged 50-60 years before and after propensity score matching (PSM)

**Figure S2:** Kaplan-Meier curves and 95% confidence intervals for death or reoperation before (A,C) and after (B,D) PSM for all patients aged 50 – 65 years (A,B) and the subgroup of patients aged 50 – 60 years (C,D)

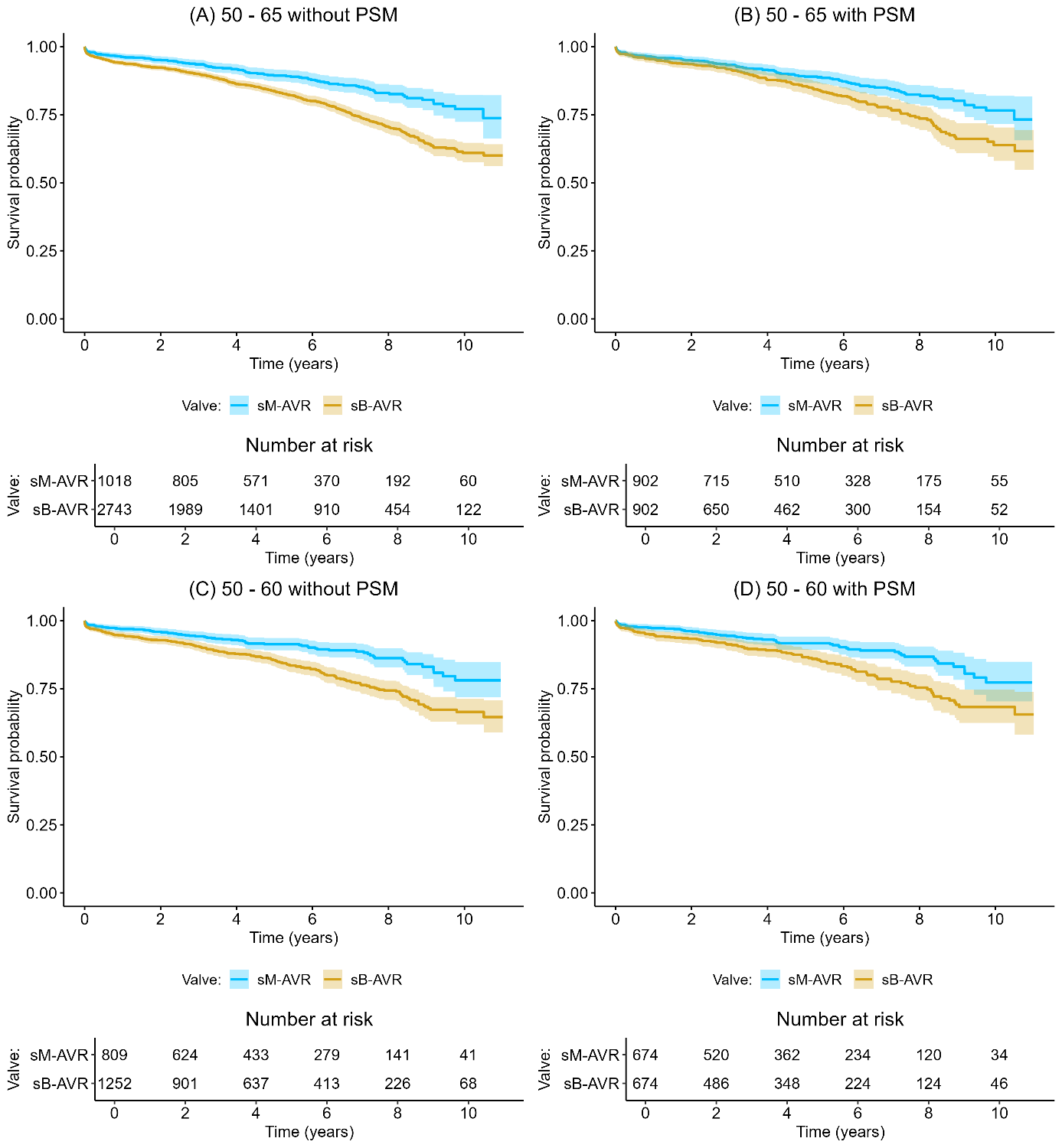
**7.3. Reoperation**

|  | Patients aged 50 – 65 years | | | | Patients aged 50 – 60 years | | | |
| --- | --- | --- | --- | --- | --- | --- | --- | --- |
|  | All data | | After PSM | | All data | | After PSM | |
| Variables | HR (95% CI) | P-value | HR (95% CI) | P-value | HR (95% CI) | P-value | HR (95% CI) | P-value |
| Original Model | | | | | | | | |
| Heart valve (sM-AVR) | 2.338  (1.360 - 4.019) | 0.002 | 2.451  (1.33 - 4.516) | 0.004 | 2.015  (1.075 - 3.778) | 0.029 | 2.579  (1.255 - 5.298) | 0.010 |
| Age | 0.994  (0.950 - 1.041) | 0.810 | 0.955  (0.889 - 1.026) | 0.210 | 0.961  (0.88 - 1.049) | 0.380 | 1.024  (0.910 - 1.152) | 0.690 |
| Sex (M) | 1.152  (0.773 - 1.716) | 0.490 | 1.616  (0.892 - 2.926) | 0.110 | 1.398  (0.779 - 2.509) | 0.260 | 1.508  (0.738 - 3.08) | 0.260 |
| Heart failure (Yes) | 1.116  (0.612 - 2.033) | 0.720 | 1.523  (0.595 - 3.902) | 0.380 | 1.386  (0.569 - 3.372) | 0.470 | 1.237  (0.36 - 4.252) | 0.740 |
| Myocardial infarction (Yes) | 0.299  (0.040 - 2.204) | 0.240 | 1.114  (0.139 - 8.922) | 0.920 | 0.712  (0.09 - 5.635) | 0.750 | 2.428  (0.269 - 21.891) | 0.430 |
| Embolic stroke or ICH (Yes) | 2.249  (0.850 - 5.951) | 0.100 | NA | NA | NA | NA | NA | NA |
| Diabetes mellitus (Yes) | 1.403  (0.866 - 2.272) | 0.170 | 1.500  (0.631 - 3.566) | 0.360 | 0.989  (0.419 - 2.336) | 0.980 | 0.744  (0.207 - 2.677) | 0.650 |
| Adiposity (Yes) | 0.836  (0.426 - 1.638) | 0.600 | 0.692  (0.202 - 2.376) | 0.560 | 0.406  (0.100 - 1.652) | 0.210 | NA | NA |
| Hyperlipidemia (Yes) | 0.778  (0.478 - 1.266) | 0.310 | 0.842  (0.399 - 1.777) | 0.650 | 0.819  (0.385 - 1.742) | 0.600 | 0.962  (0.410 - 2.256) | 0.930 |
| Hyperuricemia/gout (Yes) | 0.332  (0.046 - 2.384) | 0.270 | NA | NA | NA | NA | NA | NA |
| Valvular, rhythmological, and other CMPs (Yes) | 1.273  (0.744 - 2.178) | 0.380 | 1.492  (0.656 - 3.394) | 0.340 | 1.438  (0.675 - 3.062) | 0.350 | 1.468  (0.602 - 3.580) | 0.400 |
| Ischemic CMP (Yes) | 0.802  (0.537 - 1.200) | 0.280 | 0.828  (0.451 - 1.519) | 0.540 | 0.897  (0.487 - 1.65) | 0.730 | 0.755  (0.364 - 1.566) | 0.450 |
| Atherosclerosis (Yes) | 0.675  (0.212 - 2.151) | 0.510 | 1.797  (0.422 - 7.653) | 0.430 | 1.298  (0.293 - 5.743) | 0.730 | 1.998  (0.403 - 9.916) | 0.400 |
| Pulmonary diseases (Yes) | 1.480  (0.537 - 4.078) | 0.450 | NA | NA | 1.597  (0.373 - 6.837) | 0.530 | 1.186  (0.134 - 10.494) | 0.880 |
| Kidney diseases (Yes) | 1.094  (0.525 - 2.280) | 0.810 | 0.594  (0.131 - 2.679) | 0.500 | 0.880  (0.254 - 3.054) | 0.840 | 1.089  (0.252 - 4.700) | 0.910 |
| Malignant diseases (Yes) | 0.487  (0.117 - 2.031) | 0.320 | 1.004  (0.128 - 7.870) | 1.000 | 1.267  (0.296 - 5.423) | 0.750 | NA | NA |
| Interaction Terms | | | | | | | | |
| Heart valve (sM-AVR) × Age | 1.072  (0.947 - 1.213) | 0.270 | 1.081  (0.931 - 1.256) | 0.310 | 0.981  (0.793 - 1.213) | 0.860 | 1.040  (0.797 - 1.358) | 0.770 |
| Heart valve (sM-AVR) × Sex (M) | 1.414  (0.423 - 4.720) | 0.570 | 3.266  (0.790 - 13.507) | 0.100 | 1.353  (0.355 - 5.158) | 0.660 | 1.485  (0.318 - 6.924) | 0.610 |

**Supplementary Table 10:** Hazard ratios (HRs) and corresponding 95% confidence intervals (CIs) from multivariable competing risk regression models accounting for all listed confounders for reoperation in all patients and the subgroup of patients aged 50-60 years before and after propensity score matching (PSM)

**Figure S3:** Cumulative incidence curves for reoperation before (A,C) and after (B,D) PSM for all patients aged 50 – 65 years (A,B) and the subgroup of patients aged 50 – 60 years (C,D)

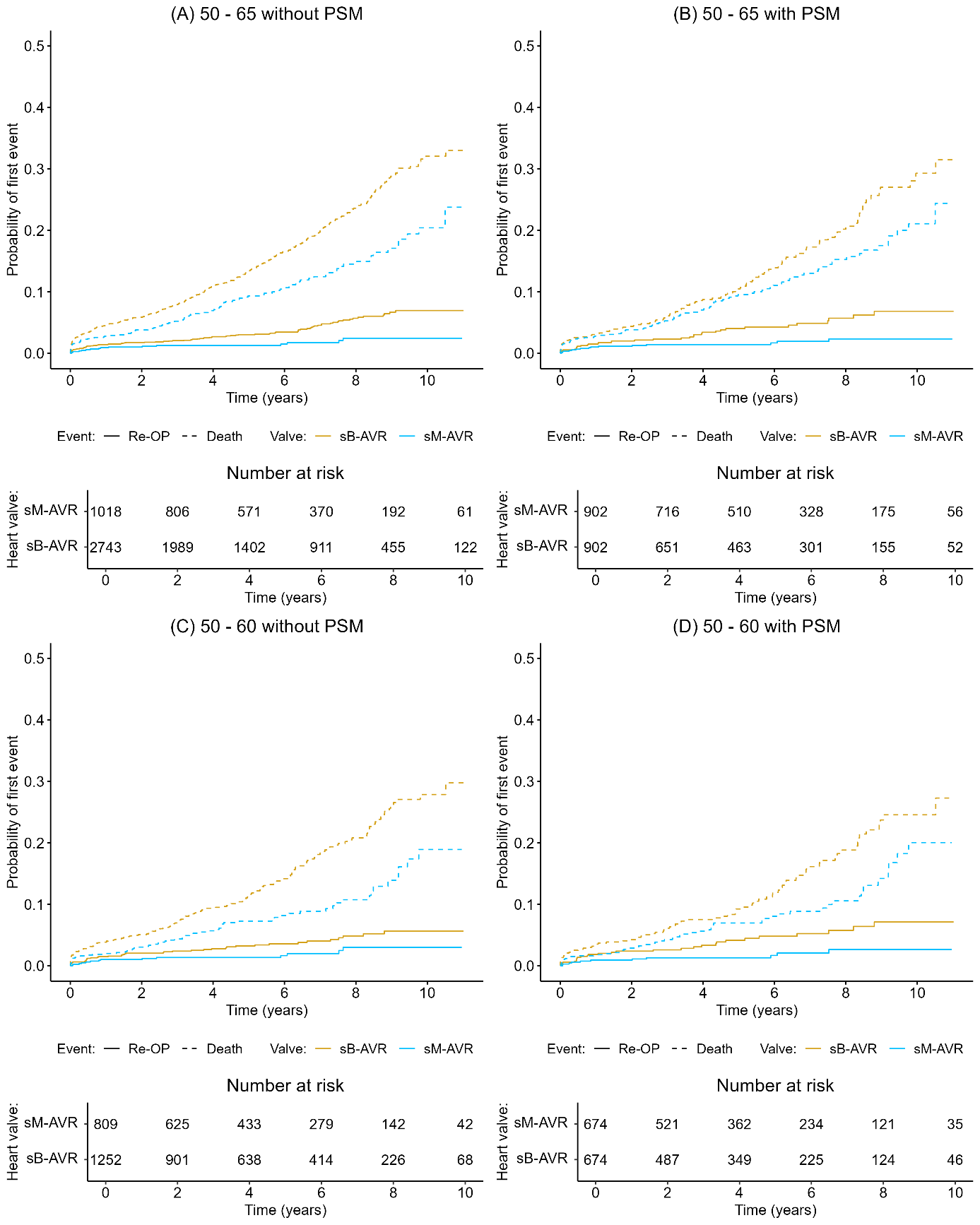

**7.4. Heart failure**

|  | Patients aged 50 – 65 years | | | | Patients aged 50 – 60 years | | | |
| --- | --- | --- | --- | --- | --- | --- | --- | --- |
|  | All data | | After PSM | | All data | | After PSM | |
| Variables | HR (95% CI) | P-value | HR (95% CI) | P-value | HR (95% CI) | P-value | HR (95% CI) | P-value |
| Original Model | | | | | | | | |
| Heart valve (sM-AVR) | 1.181  (0.909 - 1.534) | 0.210 | 1.286  (0.947 - 1.746) | 0.110 | 1.379  (0.997 - 1.908) | 0.052 | 1.457  (1.005 - 2.114) | 0.047 |
| Age | 1.040  (1.012 - 1.069) | 0.005 | 1.035  (0.997 - 1.075) | 0.072 | 1.023  (0.975 - 1.074) | 0.350 | 1.062  (0.996 - 1.133) | 0.067 |
| Sex (M) | 0.981  (0.778 - 1.237) | 0.870 | 0.867  (0.605 - 1.241) | 0.430 | 1.082  (0.764 - 1.531) | 0.660 | 0.969  (0.622 - 1.509) | 0.890 |
| Heart failure (Yes) | NA | NA | NA | NA | NA | NA | NA | NA |
| Myocardial infarction (Yes) | 1.144  (0.696 - 1.882) | 0.600 | 1.116  (0.481 - 2.586) | 0.800 | 0.896  (0.409 - 1.966) | 0.780 | 1.789  (0.653 - 4.905) | 0.260 |
| Embolic stroke or ICH (Yes) | 2.548  (1.456 - 4.459) | 0.001 | 2.596  (1.062 - 6.346) | 0.036 | 2.142  (0.975 - 4.706) | 0.058 | 1.392  (0.233 - 8.312) | 0.720 |
| Diabetes mellitus (Yes) | 1.907  (1.483 - 2.452) | <0.001 | 1.762  (1.166 - 2.664) | 0.007 | 1.886  (1.263 - 2.816) | 0.002 | 1.951  (1.163 - 3.273) | 0.011 |
| Adiposity (Yes) | 1.506  (1.107 - 2.047) | 0.009 | 1.189  (0.706 - 2.002) | 0.510 | 1.250  (0.762 - 2.05) | 0.380 | 0.629  (0.268 - 1.480) | 0.290 |
| Hyperlipidemia (Yes) | 0.734  (0.567 - 0.950) | 0.019 | 0.831  (0.570 - 1.210) | 0.330 | 1.031  (0.714 - 1.489) | 0.870 | 1.025  (0.642 - 1.637) | 0.920 |
| Hyperuricemia/gout (Yes) | 0.956  (0.508 - 1.797) | 0.89 | 0.862  (0.338 - 2.200) | 0.760 | 1.960  (0.914 - 4.206) | 0.084 | 1.679  (0.539 - 5.231) | 0.370 |
| Valvular, rhythmological, and other CMPs (Yes) | 0.780  (0.592 - 1.028) | 0.078 | 0.735  (0.492 - 1.097) | 0.130 | 0.696  (0.471 - 1.03) | 0.070 | 0.665  (0.421 - 1.049) | 0.079 |
| Ischemic CMP (Yes) | 1.086  (0.873 - 1.349) | 0.460 | 1.280  (0.920 - 1.78) | 0.140 | 1.235  (0.899 - 1.696) | 0.190 | 1.075  (0.724 - 1.596) | 0.720 |
| Atherosclerosis (Yes) | 1.630  (1.081 - 2.458) | 0.020 | 1.775  (0.909 - 3.468) | 0.093 | 1.793  (0.938 - 3.431) | 0.078 | 1.831  (0.842 - 3.98) | 0.130 |
| Pulmonary diseases (Yes) | 0.991  (0.548 - 1.794) | 0.980 | 1.022  (0.412 - 2.534) | 0.960 | 0.774  (0.275 - 2.178) | 0.630 | 0.906  (0.247 - 3.322) | 0.880 |
| Kidney diseases (Yes) | 1.997  (1.448 - 2.755) | <0.001 | 2.060  (1.254 - 3.385) | 0.004 | 2.088  (1.301 - 3.351) | 0.002 | 2.533  (1.398 - 4.590) | 0.002 |
| Malignant diseases (Yes) | 1.058  (0.587 - 1.907) | 0.850 | NA | NA | 1.612  (0.712 - 3.651) | 0.250 | NA | NA |
| Interaction Terms | | | | | | | | |
| Heart valve (sM-AVR) × Age | 0.980  (0.923 - 1.040) | 0.500 | 0.969  (0.901 - 1.041) | 0.390 | 0.995  (0.899 - 1.102) | 0.930 | 1.035  (0.913 - 1.173) | 0.590 |
| Heart valve (sM-AVR) × Sex (M) | 0.950  (0.551 - 1.637) | 0.850 | 0.839  (0.415 - 1.696) | 0.620 | 0.681  (0.335 - 1.384) | 0.290 | 0.517  (0.221 - 1.210) | 0.130 |

**Supplementary Table 11:** Hazard ratios (HRs) and corresponding 95% confidence intervals (CIs) from multivariable competing risk regression models accounting for all listed confounders for heart failure in all patients and the subgroup of patients aged 50-60 years before and after propensity score matching (PSM)

**Figure S4:** Cumulative incidence curves for heart failure before (A,C) and after (B,D) PSM for all patients aged 50 – 65 years (A,B) and the subgroup of patients aged 50 – 60 years (C,D)

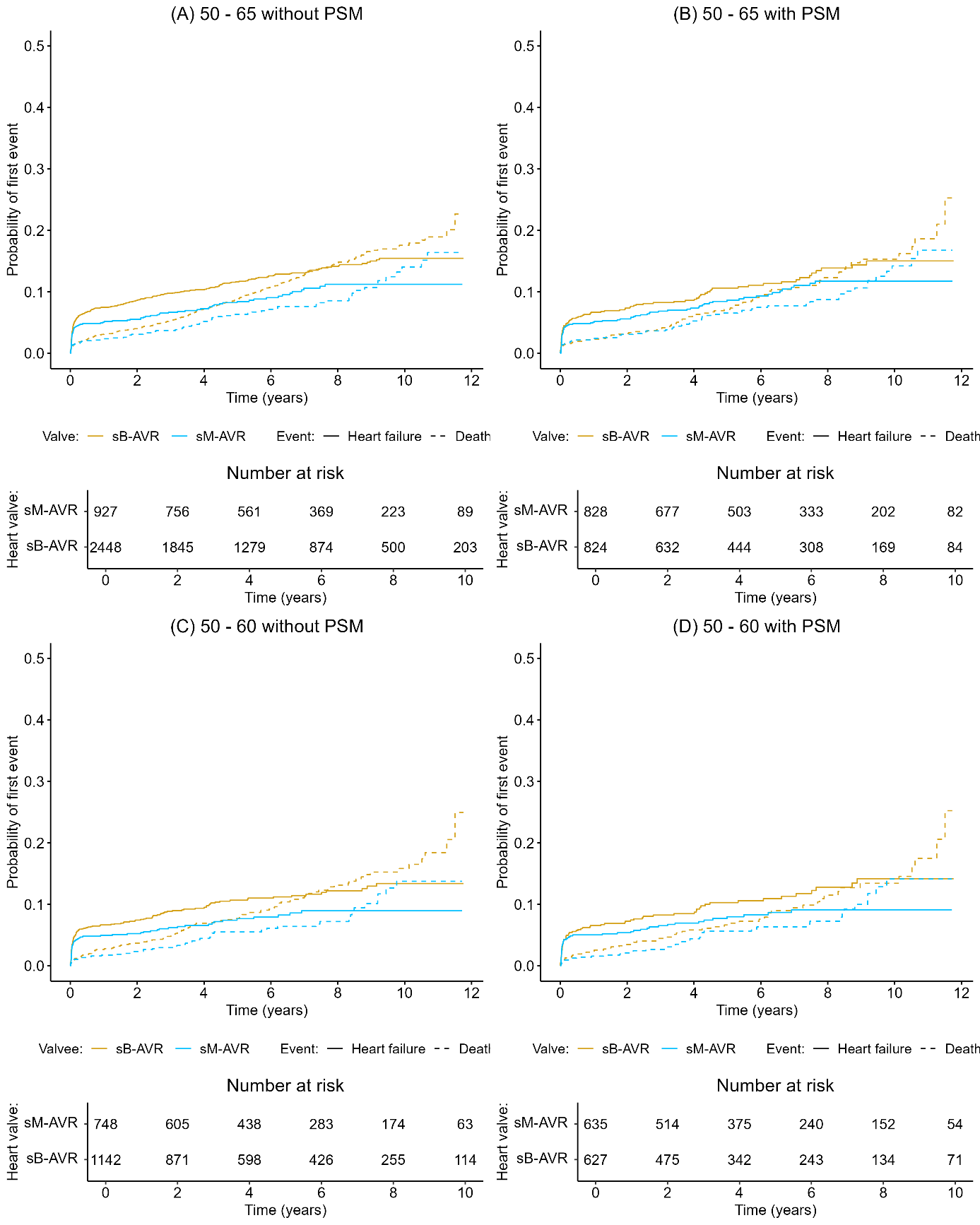

**7.5. Myocardial infarction**

|  | Patients aged 50 – 65 years | | | | Patients aged 50 – 60 years | | | |
| --- | --- | --- | --- | --- | --- | --- | --- | --- |
|  | All data | | After PSM | | All data | | After PSM | |
| Variables | HR (95% CI) | P-value | HR (95% CI) | P-value | HR (95% CI) | P-value | HR (95% CI) | P-value |
| Original Model | | | | | | | | |
| Heart valve (sM-AVR) | 1.187  (0.654 - 2.156) | 0.570 | 1.799  (0.946 - 3.42) | 0.073 | 1.421  (0.627 - 3.223) | 0.400 | 1.560  (0.634 - 3.840) | 0.330 |
| Age | 1.036  (0.970 - 1.108) | 0.290 | 1.048  (0.958 - 1.146) | 0.310 | 0.996  (0.863 - 1.149) | 0.960 | 1.021  (0.868 - 1.201) | 0.800 |
| Sex (M) | 0.968  (0.563 - 1.664) | 0.910 | 0.957  (0.484 - 1.893) | 0.900 | 0.229  (0.054 - 0.976) | 0.046 | 0.378  (0.089 - 1.607) | 0.190 |
| Heart failure (Yes) | 0.951  (0.454 - 1.992) | 0.890 | 1.391  (0.495 - 3.906) | 0.530 | 1.431  (0.443 - 4.624) | 0.550 | 0.652  (0.092 - 4.616) | 0.670 |
| Myocardial infarction (Yes) | 6.306  (3.089 - 12.873) | <0.001 | 3.487  (0.925 - 13.149) | 0.065 | 3.650  (0.984 - 13.539) | 0.053 | 2.611  (0.343 - 19.858) | 0.350 |
| Embolic stroke or ICH (Yes) | NA | NA | NA | NA | NA | NA | NA | NA |
| Diabetes mellitus (Yes) | 1.597  (0.920 - 2.773) | 0.096 | 1.624  (0.682 - 3.866) | 0.270 | 2.178  (0.847 - 5.600) | 0.110 | 2.011  (0.516 - 7.848) | 0.310 |
| Adiposity (Yes) | 1.806  (0.929 - 3.513) | 0.081 | 1.681  (0.592 - 4.770) | 0.330 | 1.743  (0.499 - 6.095) | 0.380 | 2.179  (0.359 - 13.215) | 0.400 |
| Hyperlipidemia (Yes) | 1.150  (0.672 - 1.967) | 0.610 | 1.496  (0.693 - 3.232) | 0.310 | 1.649  (0.674 - 4.030) | 0.270 | 1.624  (0.544 - 4.848) | 0.380 |
| Hyperuricemia/gout (Yes) | 1.806  (0.694 - 4.700) | 0.230 | 2.040  (0.725 - 5.741) | 0.180 | 0.932  (0.120 - 7.22) | 0.950 | 1.991  (0.296 - 13.379) | 0.480 |
| Valvular, rhythmological, and other CMPs (Yes) | 0.823  (0.415 - 1.634) | 0.580 | 0.551  (0.242 - 1.252) | 0.150 | 0.363  (0.141 - 0.932) | 0.035 | 0.278  (0.096 - 0.799) | 0.017 |
| Ischemic CMP (Yes) | 0.976  (0.567 - 1.681) | 0.930 | 1.089  (0.566 - 2.096) | 0.800 | 1.130  (0.508 - 2.516) | 0.760 | 1.452  (0.577 - 3.655) | 0.430 |
| Atherosclerosis (Yes) | 1.928  (0.849 - 4.379) | 0.120 | 3.286  (1.265 - 8.540) | 0.015 | 1.774  (0.446 - 7.05) | 0.420 | 1.401  (0.180 - 10.882) | 0.750 |
| Pulmonary diseases (Yes) | 0.404  (0.056 - 2.920) | 0.370 | 0.765  (0.099 - 5.920) | 0.800 | 1.477  (0.206 - 10.614) | 0.700 | NA | NA |
| Kidney diseases (Yes) | 0.851  (0.345 - 2.103) | 0.730 | 1.070  (0.351 - 3.268) | 0.900 | 0.499  (0.064 - 3.879) | 0.510 | 1.142  (0.157 - 8.324) | 0.900 |
| Malignant diseases (Yes) | 0.389  (0.053 - 2.876) | 0.360 | NA | NA | 1.316  (0.194 - 8.915) | 0.780 | NA | NA |
| Interaction Terms | | | | | | | | |
| Heart valve (sM-AVR) × Age | 0.912  (0.789 - 1.055) | 0.210 | 0.958  (0.809 - 1.135) | 0.620 | 0.861  (0.661 - 1.12) | 0.260 | 1.119  (0.150 - 8.349) | 0.910 |
| Heart valve (sM-AVR) × Sex (M) | 0.353  (0.111 - 1.123) | 0.078 | 0.224  (0.047 - 1.057) | 0.059 | NA | NA | NA | NA |

**Supplementary Table 12:** Hazard ratios (HRs) and corresponding 95% confidence intervals (CIs) from multivariable competing risk regression models accounting for all listed confounders for myocardial infarction in all patients and the subgroup of patients aged 50 - 60 years before and after propensity score matching (PSM)

**Figure S5:** Cumulative incidence curves for myocardial infarction before (A,C) and after (B,D) PSM for all patients aged 50 – 65 years (A,B) and the subgroup of patients aged 50 – 60 years (C,D)

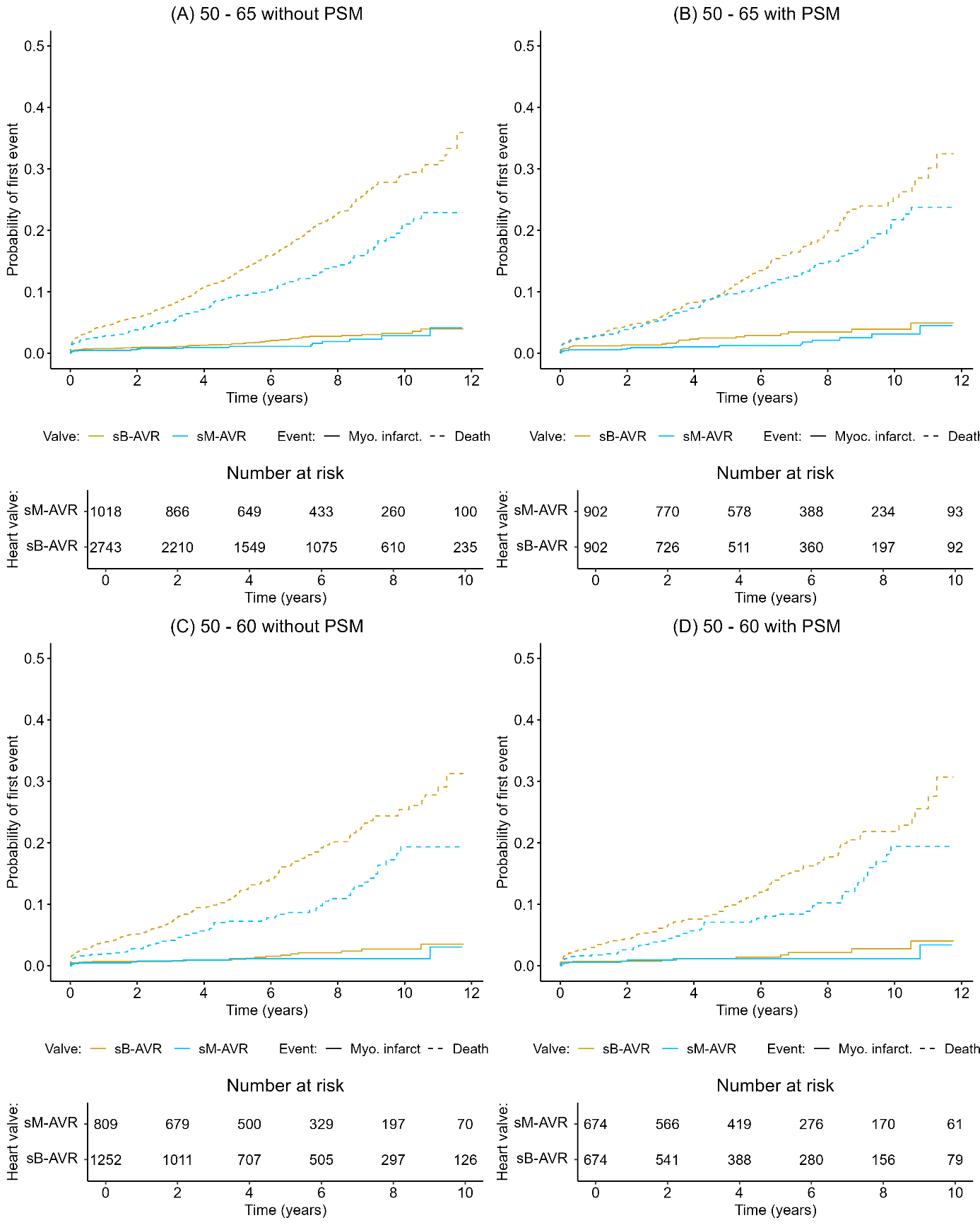

**7.6. Embolic stroke or ICH**

|  | Patients aged 50 – 65 years | | | | Patients aged 50 – 60 years | | | |
| --- | --- | --- | --- | --- | --- | --- | --- | --- |
|  | All data | | After PSM | | All data | | After PSM | |
| Variables | HR (95% CI) | P-value | HR (95% CI) | P-value | HR (95% CI) | P-value | HR (95% CI) | P-value |
| Original Model | | | | | | | | |
| Heart valve (sM-AVR) | 1.139  (0.809 - 1.604) | 0.460 | 1.209  (0.820 - 1.783) | 0.340 | 1.362  (0.907 - 2.045) | 0.140 | 1.516  (0.980 - 2.344) | 0.062 |
| Age | 1.003  (0.970 - 1.038) | 0.860 | 1.024  (0.980 - 1.070) | 0.290 | 1.036  (0.974 - 1.102) | 0.260 | 1.077  (0.997 - 1.163) | 0.060 |
| Sex (M) | 1.127  (0.831 - 1.529) | 0.440 | 0.901  (0.564 - 1.439) | 0.660 | 1.186  (0.783 - 1.797) | 0.420 | 1.028  (0.610 - 1.732) | 0.920 |
| Heart failure (Yes) | 0.617  (0.368 - 1.037) | 0.068 | 0.458  (0.188 - 1.115) | 0.085 | 0.587  (0.267 - 1.293) | 0.190 | 0.463  (0.141 - 1.519) | 0.200 |
| Myocardial infarction (Yes) | 0.696  (0.281 - 1.720) | 0.430 | 0.325  (0.040 - 2.656) | 0.290 | 0.473  (0.103 - 2.159) | 0.330 | 0.774  (0.096 - 6.254) | 0.810 |
| Embolic stroke or ICH (Yes) | 3.033  (1.474 - 6.238) | 0.003 | 3.719  (1.157 - 11.953) | 0.027 | 2.918  (1.068 - 7.969) | 0.037 | 2.577  (0.283 - 23.474) | 0.400 |
| Diabetes mellitus (Yes) | 1.476  (1.048 - 2.079) | 0.026 | 1.569  (0.943 - 2.613) | 0.083 | 1.659  (1.018 - 2.703) | 0.042 | 1.513  (0.795 - 2.878) | 0.210 |
| Adiposity (Yes) | 1.458  (0.965 - 2.205) | 0.074 | 0.850  (0.407 - 1.778) | 0.670 | 1.470  (0.812 - 2.66) | 0.200 | 1.251  (0.554 - 2.829) | 0.590 |
| Hyperlipidemia (Yes) | 1.130  (0.815 - 1.568) | 0.460 | 1.421  (0.885 - 2.281) | 0.150 | 1.142  (0.729 - 1.788) | 0.560 | 1.233  (0.708 - 2.149) | 0.460 |
| Hyperuricemia/gout (Yes) | 1.248  (0.609 - 2.556) | 0.550 | 0.655  (0.212 - 2.023) | 0.460 | 0.336  (0.048 - 2.376) | 0.270 | 0.452  (0.065 - 3.124) | 0.420 |
| Valvular, rhythmological, and other CMPs (Yes) | 0.953  (0.648 - 1.404) | 0.810 | 1.034  (0.599 - 1.786) | 0.900 | 0.938  (0.557 - 1.579) | 0.810 | 1.131  (0.609 - 2.100) | 0.700 |
| Ischemic CMP (Yes) | 0.954  (0.705 - 1.291) | 0.760 | 0.888  (0.578 - 1.363) | 0.590 | 1.087  (0.726 - 1.626) | 0.690 | 0.948  (0.587 - 1.530) | 0.830 |
| Atherosclerosis (Yes) | 1.063  (0.564 - 2.004) | 0.850 | 1.221  (0.459 - 3.252) | 0.690 | 0.454  (0.109 - 1.886) | 0.280 | 0.709  (0.164 - 3.066) | 0.640 |
| Pulmonary diseases (Yes) | 1.488  (0.728 - 3.042) | 0.280 | 2.710  (1.162 - 6.32) | 0.021 | 1.997  (0.812 - 4.909) | 0.130 | 1.854  (0.595 - 5.780) | 0.290 |
| Kidney diseases (Yes) | 1.022  (0.585 - 1.787) | 0.940 | 1.234  (0.549 - 2.77) | 0.610 | 1.386  (0.676 - 2.843) | 0.370 | 1.573  (0.637 - 3.886) | 0.330 |
| Malignant diseases (Yes) | 0.434  (0.135 - 1.397) | 0.160 | 0.578  (0.077 - 4.349) | 0.590 | 0.567  (0.130 - 2.478) | 0.450 | NA | NA |
| Interaction Terms | | | | | | | | |
| Heart valve (sM-AVR) × Age | 0.905  (0.846 - 0.968) | 0.004 | 0.907  (0.835 - 0.985) | 0.021 | 0.871  (0.769 - 0.986) | 0.029 | 0.934  (0.809 - 1.078) | 0.350 |
| Heart valve (sM-AVR) × Sex (M) | 1.012  (0.513 - 1.995) | 0.970 | 0.958  (0.398 - 2.307) | 0.920 | 1.157  (0.483 - 2.770) | 0.740 | 0.646  (0.241 - 1.731) | 0.380 |

**Supplementary Table 13:** Hazard ratios (HRs) and corresponding 95% confidence intervals (CIs) from multivariable competing risk regression models accounting for all listed confounders for embolic stroke or intracerebral hemorrhage (ICH) in all patients and the subgroup of patients aged 50-60 years before and after propensity score matching (PSM)

**Figure S6:** Cumulative incidence curves for embolic stroke or ICH before (A,C) and after (B,D) PSM for all patients aged 50 – 65 years (A,B) and the subgroup of patients aged 50 – 60 years (C,D)

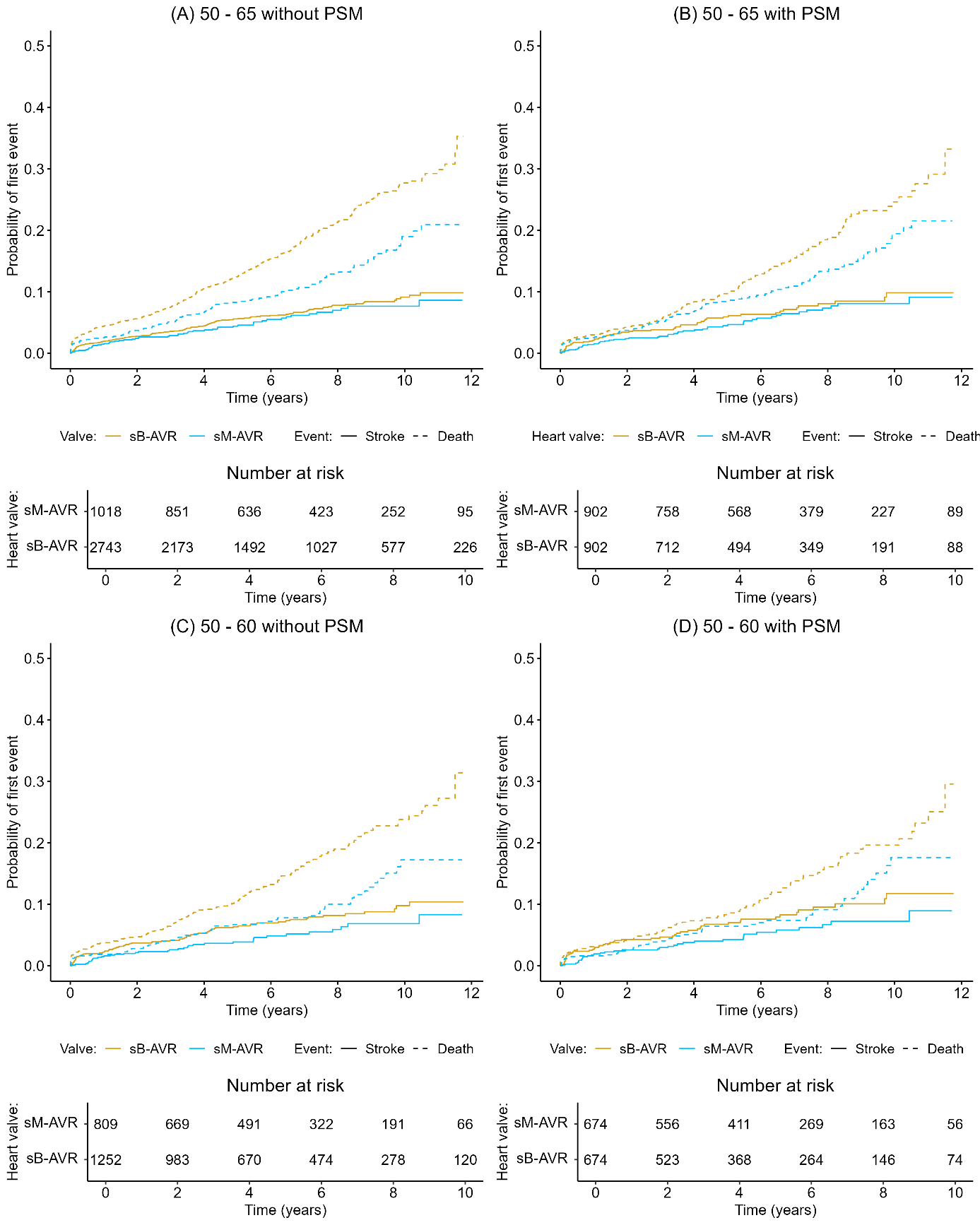

**7.7. Bleeding other than embolic stroke or intracerebral hemorrhage**

|  | Patients aged 50 – 65 years | | | | Patients aged 50 – 60 years | | | |
| --- | --- | --- | --- | --- | --- | --- | --- | --- |
|  | All data | | After PSM | | All data | | After PSM | |
| Variables | HR (95% CI) | P-value | HR (95% CI) | P-value | HR (95% CI) | P-value | HR (95% CI) | P-value |
| Original Model | | | | | | | | |
| Heart valve (sM-AVR) | 0.819  (0.561 - 1.196) | 0.300 | 0.697  (0.437 - 1.111) | 0.130 | 0.825  (0.516 - 1.320) | 0.420 | 0.676  (0.387 - 1.183) | 0.170 |
| Age | 0.991  (0.952 - 1.033) | 0.670 | 0.999  (0.942 - 1.058) | 0.960 | 0.979  (0.911 - 1.051) | 0.550 | 1.008  (0.917 - 1.109) | 0.870 |
| Sex (M) | 1.207  (0.842 - 1.730) | 0.310 | 1.015  (0.599 - 1.721) | 0.960 | 1.263  (0.766 - 2.081) | 0.360 | 1.188  (0.653 - 2.160) | 0.570 |
| Heart failure (Yes) | 1.068  (0.653 - 1.748) | 0.790 | 0.690  (0.275 - 1.732) | 0.430 | 0.884  (0.396 - 1.972) | 0.760 | 0.273  (0.036 - 2.055) | 0.210 |
| Myocardial infarction (Yes) | 0.819  (0.328 - 2.045) | 0.670 | 0.775  (0.197 - 3.044) | 0.720 | 0.672  (0.157 - 2.868) | 0.590 | 0.925  (0.155 - 5.535) | 0.930 |
| Embolic stroke or ICH (Yes) | 2.434  (0.950 - 6.237) | 0.064 | NA | NA | 2.764  (0.800 - 9.552) | 0.110 | NA | NA |
| Diabetes mellitus (Yes) | 1.385  (0.922 - 2.081) | 0.120 | 1.514  (0.810 - 2.831) | 0.190 | 1.012  (0.527 - 1.941) | 0.970 | 0.964  (0.387 - 2.402) | 0.940 |
| Adiposity (Yes) | 0.858  (0.467 - 1.575) | 0.620 | 0.600  (0.208 - 1.728) | 0.340 | 0.724  (0.290 - 1.812) | 0.490 | 0.486  (0.108 - 2.176) | 0.350 |
| Hyperlipidemia (Yes) | 0.881  (0.594 - 1.308) | 0.530 | 1.120  (0.645 - 1.945) | 0.690 | 1.113  (0.657 - 1.886) | 0.690 | 1.828  (0.990 - 3.375) | 0.054 |
| Hyperuricemia/gout (Yes) | 1.913  (0.936 - 3.906) | 0.075 | 1.071  (0.313 - 3.667) | 0.910 | 1.867  (0.640 - 5.448) | 0.250 | NA | NA |
| Valvular, rhythmological, and other CMPs (Yes) | 1.115  (0.689 - 1.804) | 0.660 | 1.195  (0.596 - 2.395) | 0.620 | 1.411  (0.707 - 2.816) | 0.330 | 1.076  (0.505 - 2.292) | 0.850 |
| Ischemic CMP (Yes) | 1.265  (0.910 - 1.758) | 0.160 | 1.845  (1.179 - 2.889) | 0.007 | 1.452  (0.932 - 2.264) | 0.099 | 1.214  (0.707 - 2.085) | 0.480 |
| Atherosclerosis (Yes) | 1.820  (1.023 - 3.237) | 0.042 | 2.511  (1.129 - 5.586) | 0.024 | 1.383  (0.512 - 3.733) | 0.520 | 1.876  (0.669 - 5.261) | 0.230 |
| Pulmonary diseases (Yes) | 0.611  (0.191 - 1.952) | 0.410 | NA | NA | NA | NA | NA | NA |
| Kidney diseases (Yes) | 1.552  (0.915 - 2.632) | 0.100 | 0.898  (0.350 - 2.303) | 0.820 | 1.998  (0.999 - 3.994) | 0.050 | 1.619  (0.639 - 4.104) | 0.310 |
| Malignant diseases (Yes) | 0.939  (0.388 - 2.270) | 0.890 | 2.029  (0.692 - 5.947) | 0.200 | 0.869  (0.228 - 3.308) | 0.840 | NA | NA |
| Interaction Terms | | | | | | | | |
| Heart valve (sM-AVR) × Age | 0.961  (0.885 - 1.043) | 0.340 | 0.954  (0.851 - 1.070) | 0.420 | 0.884  (0.764 - 1.023) | 0.098 | 0.861  (0.718 - 1.034) | 0.110 |
| Heart valve (sM-AVR) × Sex (M) | 2.439  (1.054 - 5.641) | 0.037 | 2.995  (1.021 - 8.792) | 0.046 | 2.892  (1.026 - 8.149) | 0.045 | 2.491  (0.746 - 8.314) | 0.140 |

**Supplementary Table 14:** Hazard ratios (HRs) and corresponding 95% confidence intervals (CIs) from multivariable competing risk regression models accounting for all listed confounders for bleeding other than embolic stroke or intracerebral hemorrhage (ICH) in all patients and the subgroup of patients aged 50 - 60 years before and after propensity score matching (PSM)

**Figure S7:** Cumulative incidence curves for bleeding other than embolic stroke or ICH before (A,C) and after (B,D) PSM for all patients aged 50 – 65 years (A,B) and the subgroup of patients aged 50 – 60 years (C,D)

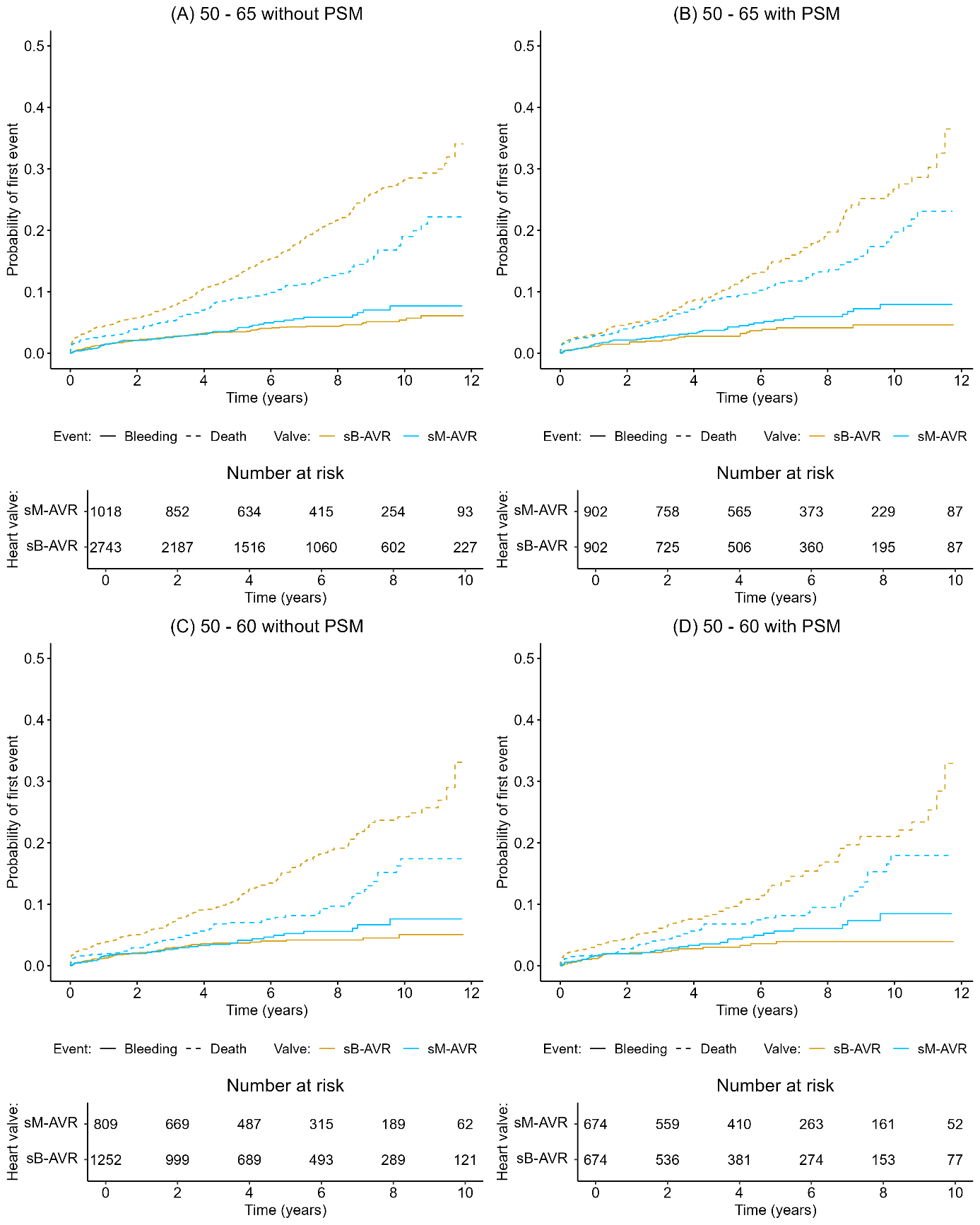

**8.) Exploratory Outcome**

**Figure S8:** Kaplan-Meier curves and 95% confidence intervals for survival after reoperation for all patients aged 50 – 65 years (A) and the subgroup of patients aged 50 – 60 years (B)

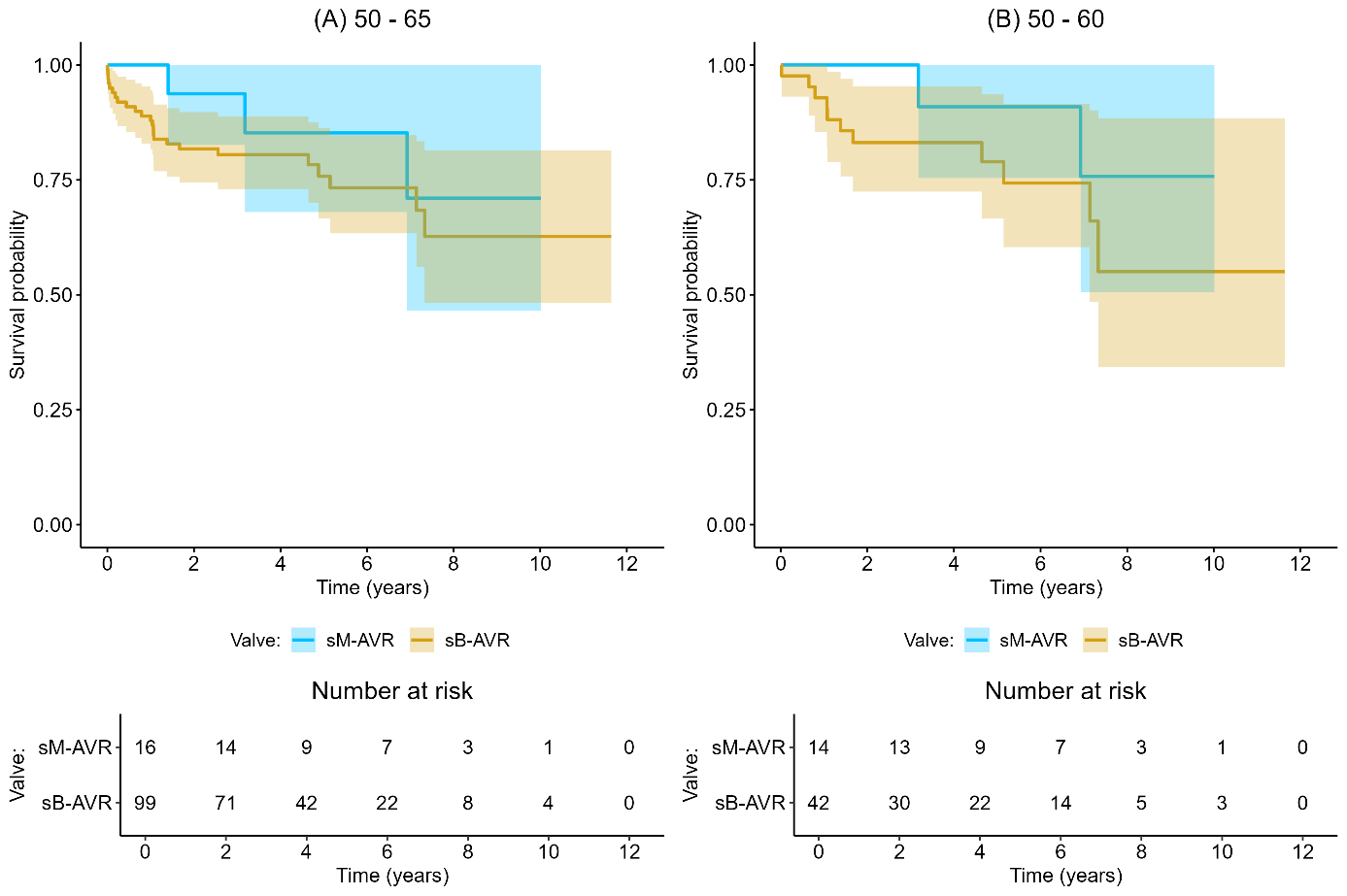
